# Impact of oral cholera vaccine outbreak response immunization in sub-Saharan Africa, 2010–2020: a modeling analysis

**DOI:** 10.1101/2025.11.06.25339715

**Authors:** Jong-Hoon Kim, Monica Duong, Elizabeth C. Lee, Qulu Zheng, Andrew S. Azman, Allyson Russell, Jerome H. Kim

**Author notes:** Correspondence: Jong-Hoon Kim.

## Abstract

**Background:** Oral cholera vaccine (OCV) is a critical tool for controlling cholera outbreaks. However, the effectiveness and efficiency of reactive OCV campaigns depend strongly on implementation timing and targeting strategy.

**Methods:** Using data from 1,192 cholera outbreaks (2010–2020) in sub-Saharan Africa, we conducted stochastic simulations of single-dose OCV outbreak response immunization (ORI) strategies varying by timing, coverage, triggering criteria, and outbreak selection. Outcomes included impact (cases, deaths, and DALYs averted), efficiency (impact per 1,000 OCV doses), and cost-effectiveness (cost per unit of impact).

**Findings:** Week-1 ORIs with 75% vaccine coverage averted a median of 70% of cases (129 cases, IQR 41–375) whereas week-9 ORIs—comparable to historical response times—averted only ∼8% (16 cases; IQR 0–112). Effectiveness declined exponentially by 23.6% per week (IQR 10.7–77.4), halving every 2.6 weeks. Efficiency fell from 0.69 to 0.09 cases averted per 1,000 doses, and cost-effectiveness worsened from US$4,662 to US$13,043 per case averted. When ORIs were triggered after case accumulation, effectiveness decreased by 6.4% (IQR 2.5–4.2) for every 10 additional cases, halving every ∼104 cases. Prioritizing large or high-attack-rate outbreaks (top 10%) improved efficiency 14–27-fold and reduced cost per DALY averted by 95%–99%. Even with delays, week-9 ORIs deployed across multiple outbreaks, achieved a median 0.44 (IQR: 0.28–0.44) cases averted per 1,000 doses, which increasing further with targeted deployment.

**Interpretation:** Rapid deployment of OCV, with priority to large and high-attack-rate outbreaks, is essential to maximize the impact, efficiency, and cost-effectiveness of cholera response in Africa. Even delayed reactive vaccination can meaningfully reduce cholera burden.

**Key messages:**

1. We analyzed 1,192 cholera outbreaks across sub-Saharan Africa (2010–2020) to estimate the impact, efficiency, and cost effectiveness of single-dose oral cholera vaccine (OCV) outbreak response immunization (ORIs).
2. ORI implemented in week 1 of outbreak averted a median 129 cases (IQR 41–375) averted, corresponding to 70% (IQR 49.8–82.5) reduction; impact declined sharply with delay, halving each median 2.6 (IQR 0.5–6.1) weeks.
3. Prioritizing large, prolonged, and high-attack-rate outbreaks markedly improved impact, efficiency, and cost-effectiveness, with up to 99% reduction in cost per DALY averted among the top 10% of outbreaks by attack rate.
4. In simulations reflecting historical OCV stockpile use, even delayed week-9 ORIs averted 0·44 (IQR 0·28–0·44) cases per 1,000 doses, increasing further with targeted deployment.
5. Rapid and targeted deployment of reactive OCV is essential to maximize the impact of limited global OCV supply and to reduce cholera burden in Africa.

**Research in context:** *Evidence before this study:* We searched PubMed on March 15, 2024, without language or date restrictions, using the search terms ((“Cholera”[Mesh] OR “Vibrio cholerae”[Mesh])) AND “Cholera Vaccines”[Mesh] AND ((model*[Title/Abstract]) OR (math*[Title/Abstract])) AND (vaccine*[Title/Abstract]). The search yielded 164 articles, of which 17 studies met the inclusion criteria, focusing on mathematical models assessing the impact of oral cholera vaccines (OCVs) under different scenarios. Most studies simulated outbreaks in single geographic settings—such as Bangladesh^1–4^, Chad^5^, Haiti^6–8^, South Sudan,^9^ or Zimbabwe^10^— or not anchored in a specific setting.^11^ A few analyses considered outbreaks across multiple countries.^12–16^ These studies varied widely in campaign design (reactive mass campaigns, proactive campaigns, “hotspot” or area-targeted campaigns), outbreak scale, and modeling assumptions, including delays in vaccine delivery and onset of protection. While some models assumed protection immediately following vaccination^13,14,17,18^, others considered a lag between vaccination and protection.^5,7,12,19^ Most previous work focused on large, well-documented outbreaks (e.g., Zimbabwe with 98,591 cases or Haiti with 119,902 cases)^14,19^ and did not represent the outbreaks that may be typically found in sub-Saharan Africa. These gaps highlight the need for models that capture heterogeneity of outbreaks to improve the generalizability of vaccine impact assessments.

*Added value of this study:* To our knowledge, this is the first study to model the impact, efficiency, and cost-effectiveness of OCV outbreak response immunizations (ORIs) across a large and diverse set of outbreaks from multiple temporal and geographic contexts in sub-Saharan Africa. Using data from 1,192 outbreaks from 2010 to 2020—spanning wide variation in size, duration, and attack rate—we developed a comprehensive framework to evaluate how vaccination timing, coverage, and targeting influence outcomes. Unlike previous studies focused on isolated large outbreaks, our model quantifies the distribution of vaccine impact across heterogeneous epidemics, providing a generalizable basis for reactive vaccination planning and optimization of global OCV stockpile use.

*Implications of the available evidence:* Our findings underscore the critical importance of rapid and targeted vaccine deployment. ORI impact declined by half approximately every 2·6 weeks of implementation delay, yet substantial benefits persisted even with late campaigns, particularly in large and high–attack-rate outbreaks. When aggregated across outbreaks, week-9 ORIs, comparable to historical response times, still averted 0·44 [IQR: 0·28–0·44] cases per 1,000 OCV doses, rising to 2·2 [IQR: 1·9–2·2] with week-1 campaigns targeting the top 70% of outbreaks by attack rate. Prioritizing large, sustained outbreaks increased cost-effectiveness of week-9 ORI by up to 85-fold. These results provide a quantitative basis for optimizing reactive vaccination strategies under constrained OCV supply to maximize health impact across Africa.

## Introduction

Cholera, one of the oldest infectious diseases affecting humans, has persisted as a major public health challenge since the 7th pandemic began near the Bay of Bengal in 1961.^20^ It remains a significant threat in settings with inadequate water, sanitation, and hygiene (WaSH), particularly in low- and middle-income countries (LMICs).^21^ Modelled estimates and World Health Organization (WHO) reports indicate that sub-Saharan Africa bears most of the global burden, marked by substantial spatial heterogeneity.^22–24^ An analysis of cholera incidence in the region from 2010 to 2016 identified nearly 1,000 outbreaks of varying scale across multiple administrative levels,^25^ underscoring the complexity of controlling transmission.

Sustained access to adequate WaSH—together with case management and surveillance— remains the foundation of cholera control. However, oral cholera vaccines (OCVs) provide a critical complementary tool, particularly where long-term infrastructure improvements are difficult to achieve. To facilitate rapid vaccine access, WHO established the global OCV stockpile in 2013 with Gavi’s support.^26^ Between 2013 and 2024, more than 200 million doses have been deployed for both proactive and reactive campaigns.^27^ In response to growing demand and limited supply, the International Coordinating Group (ICG)—which manages the stockpile—temporarily adopted a single-dose regimen for outbreak response in October 2022, replacing the standard two-dose schedule.^26^

Given constrained vaccine supply, optimizing the use of available doses is critical. While preemptive vaccination—targeting high-risk areas identified through environmental or historical risk indicators^28^, or enhanced diagnostics^16^—can be effective, such approaches are challenged by fluctuating incidence,^29^ waning immunity,^30^ and population mobility.^17,31^ In contrast, outbreak response immunizations (ORIs), which vaccinate populations in locations experiencing ongoing transmission, avoid these uncertainties. In practice, more than 75% of over 200 million doses deployed by 2024 were used for ORIs rather than preventive campaigns.^27^

Despite this widespread use, evidence on ORI impact remains fragmented. Most previous analyses have focused a limited number of large outbreaks,^2,5–7,12,14,25^ which do not represent the diverse epidemiological profiles of cholera outbreaks,^25,32^ or have overlooked key operational and immunological factors such ss delayed campaign initiation^27,33^ or time required to develop vaccine-derived immunity.^13,14,17,18^ A comprehensive modeling framework that integrates these factors an captures the epidemiological diversity of real-world outbreaks is therefore essential to guide evidence-based vaccine policy.

We addressed this evidence gaps by evaluating the health and economic impact of single-dose OCV ORIs across 1,192 cholera outbreaks in sub-Saharan Africa between 2010 and 2020. Specifically, we asked: (1) how effective are single-dose OCV ORIs when implemented under varying delays and coverage levels during individual outbreaks? and (2) how effective are ORIs when deployed across multiple heterogeneous outbreaks, as in global stockpile operations? To answer these questions, we combined outbreak-level impact estimates with aggregate analyses simulating vaccine deployment under stockpile constraints across a range of operational and prioritization scenarios.

## Methods

### Outbreak data

We compiled weekly suspected cholera incidence data from the Cholera Taxonomy Data,^34^ which includes 1,664 outbreaks reported across various administrative levels in 30 countries in sub-Saharan Africa from January 2010 to July 2023.^25,35^ Outbreaks were defined by location-specific average weekly incidence thresholds, spanning from the first to the last reported suspected case, with weeks without data assumed to have zero cases.^32^

To ensure consistency with real-world outbreak response conditions, several exclusion criteria were applied. Outbreaks were excluded if they occurred in administrative units whose population sizes fell outside the range of historical OCV deployments (i.e., between 16,500 and 5 million doses).^36^ When outbreaks overlapped across multiple administrative levels, smaller outbreaks contained within a larger one were excluded. If a smaller administrative unit reported an equal or greater number of cases, the outbreak from the larger administrative unit was excluded. Outbreaks that occurred within three years of a prior OCV campaign were excluded to avoid vaccine-influenced incidence patterns that could bias counterfactual analyses. Finally, outbreaks beginning after Jan 1, 2021 were excluded due to incomplete OCV campaign records. After applying these criteria, 1,192 outbreaks were retained for modeling analyses. (**Figure 1A**).

**Figure 1.**
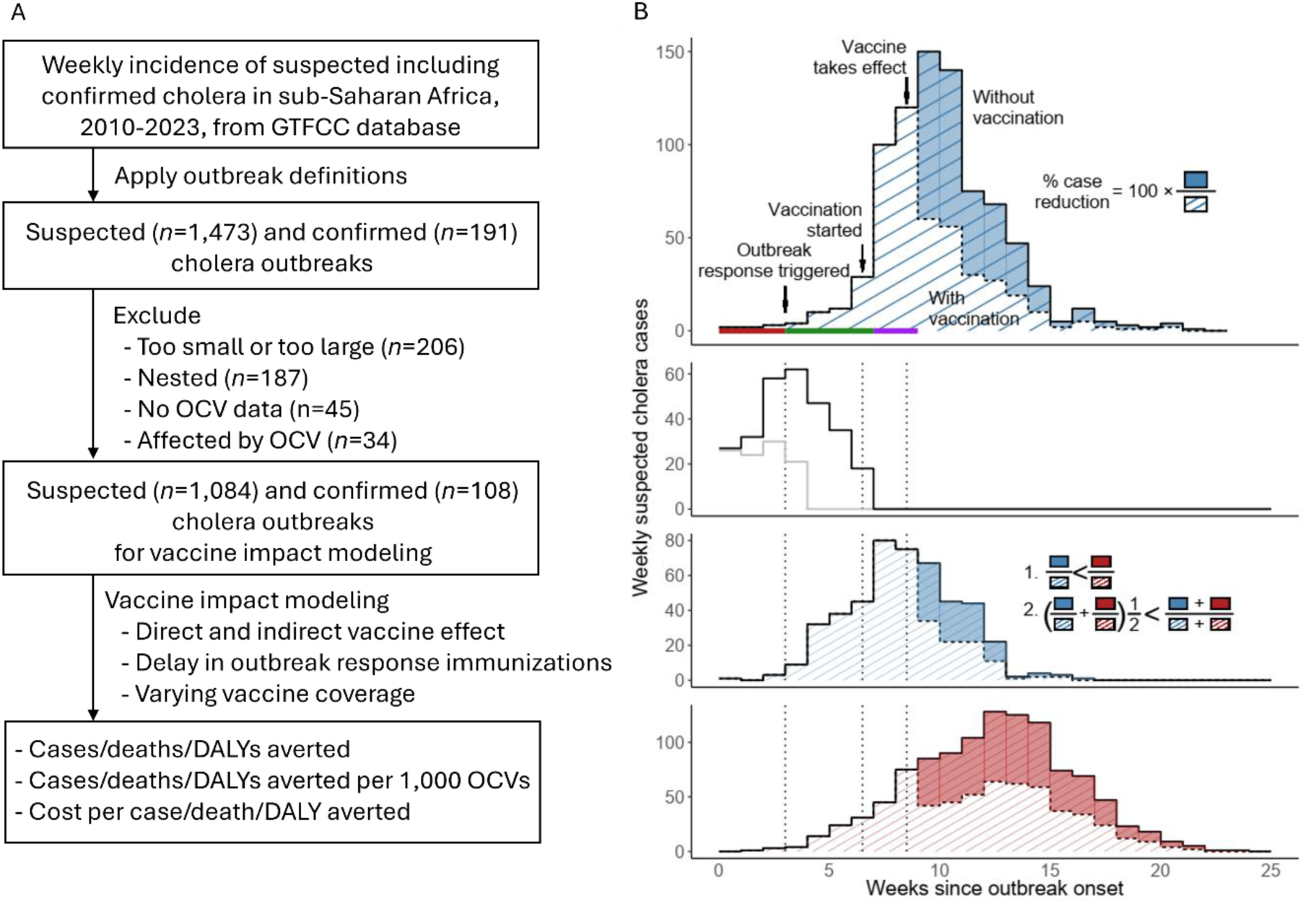
Overview of outbreak identification process and modeling of vaccination impact. (A) Flowchart showing the identification of cholera outbreaks and selecting a subset for vaccine impact modeling. (B) Schematic illustrating vaccine impact under different outbreak scenarios. The top panel shows a hypothetical cholera outbreak with and without vaccination. Outbreak response can be triggered either at outbreak onset or by reaching a cumulative case threshold (red thick line). Vaccination then occurs after a random delay (thick green line) and vaccines take effect again after a random delay following vaccination (thick purple line). Percentage reduction in cases is calculated as the ratio of cases reduced because of vaccines to cases under no vaccination. The second panel demonstrates early outbreaks that end before vaccination takes place (grey line) or before vaccines effectively protect vaccine recipients and others (black line). The third and the bottom panel show that vaccination impact in a smaller and a larger outbreak, respectively. The percentage reduction in cases is typically larger in outbreaks with more cases (inequality 1 in the third panel). The overall percentage reduction in cases across multiple heterogeneous outbreaks is larger when outbreaks are weighted by their total case count than when each outbreak is given equal weight regardless of size (inequality 2 in the third panel).

### Modeling the vaccine impact

We simulated weekly case counts under counterfactual vaccination scenarios—identical in all respects except for the implementation of ORIs. The impact of ORIs was based on the number of cases averted through vaccination (**Figure 1B**, top panel). Because ORIs can only occur if outbreaks are still ongoing, we assumed no campaign would be initiated if two or more weeks had passed since the outbreak ended, consistent with the outbreak definition^37^ and validated in sensitivity analyses. ORIs beginning before outbreak resolution may still produce no impact due to the time required for implementation and vaccine-derived immunity to develop (**Figure 1B**, second panel). The impact of ORIs tends to be larger for a larger outbreak (**Figure 1B**, third panel) and the overall impact across multiple heterogeneous outbreaks tends to be larger than the simple average, as larger outbreaks contribute disproportionately to the overall impact (**Figure 1B**, fourth panel).

The model captures both direct and indirect effects of OCV^38^ (Supplementary Information **S1**) and incorporates age-specific vaccine efficacy (< 5 and 5+ yo), delays in ORI implementation, and the delays in the vaccine-derived immunity (**Table 1**). The proportion of cases averted, *P*_*i*,*π*_, at a given vaccine coverage *π* for each age group—children under five (*i* = 1) and individuals aged five or older (*i* = 2)—was computed as:

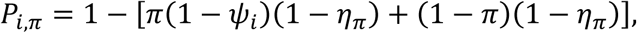

where *ψ*_*i*_represents direct vaccine efficacy for age group *i* and *η*_*π*_ denotes the indirect vaccine efficacy at coverage *π*. Thus, unvaccinated individuals, comprising 1 − *π* of the population, still benefit from indirect protection 1 − *η*_*π*_. We estimated *η*_*π*_from reduction in incidence rates among unvaccinated individuals observed in a randomized trial conducted in Matlab, Bangladesh using Bayesian beta regression.^1,39^ (Supplementary Information **S2** and **Figure S1**). The cumulative number of cases, *C*_*i*,*π*,*τ*_, for age group *i*, for a vaccination campaign initiated at time *τ* with coverage *π* was calculated as:

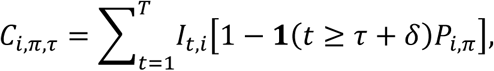

where *I*_*t*,*i*,_is the incidence at time *t* for age group *i* and **1**(*x*) is an indicator function (1 if *x* is true, 0 otherwise), *δ* captures delays for ORI implementation and the vaccine-derived immunity, and *T* marks the end of the outbreak. Overall, we simulated ORIs in each of the 1,192 outbreaks under combinations of vaccination coverage, campaign timing, and trigger criteria (outbreak onset-triggered or case-triggered) (**Table 2**). Two hundred parameter sets were drawn using a Sobol low-discrepancy sequence (**Table 3** and Supplementary Information **S3**) and results are summarized as medians with interquartile ranges (IQRs). The outcomes were reported as *impact*, i.e., cases, deaths, and disability-adjusted life years (DALYs) averted, which were also converted to *efficiency* (i.e., impact per 1,000 OCVs) and *cost-effectiveness* metrics (e.g., cost per unit of impact). All code and processed datasets are publicly available in a GitHub repository.^40^

**Table 1.**
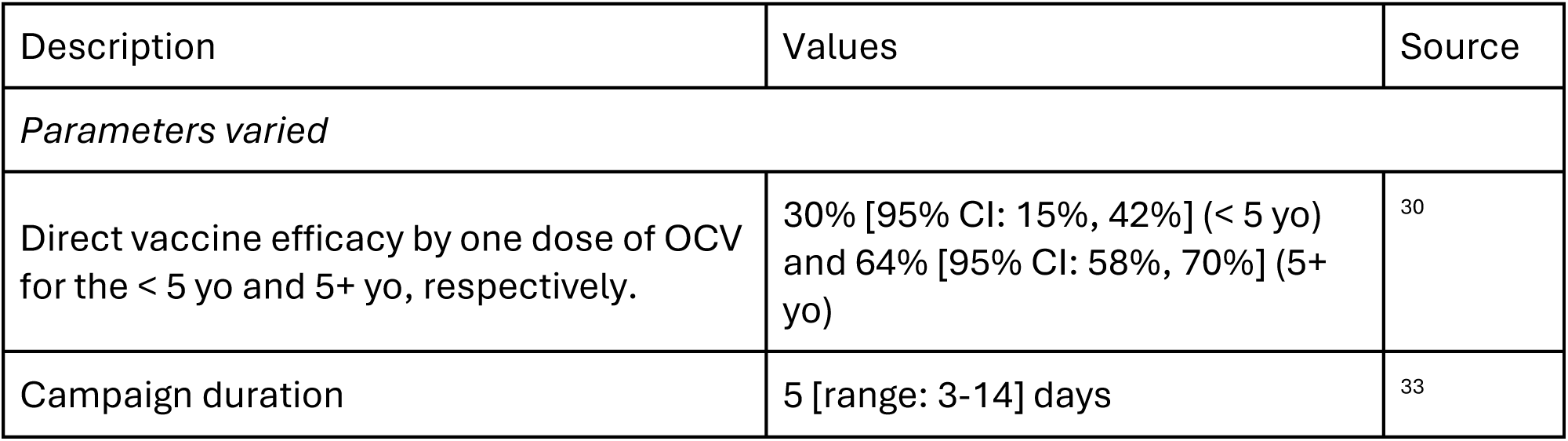
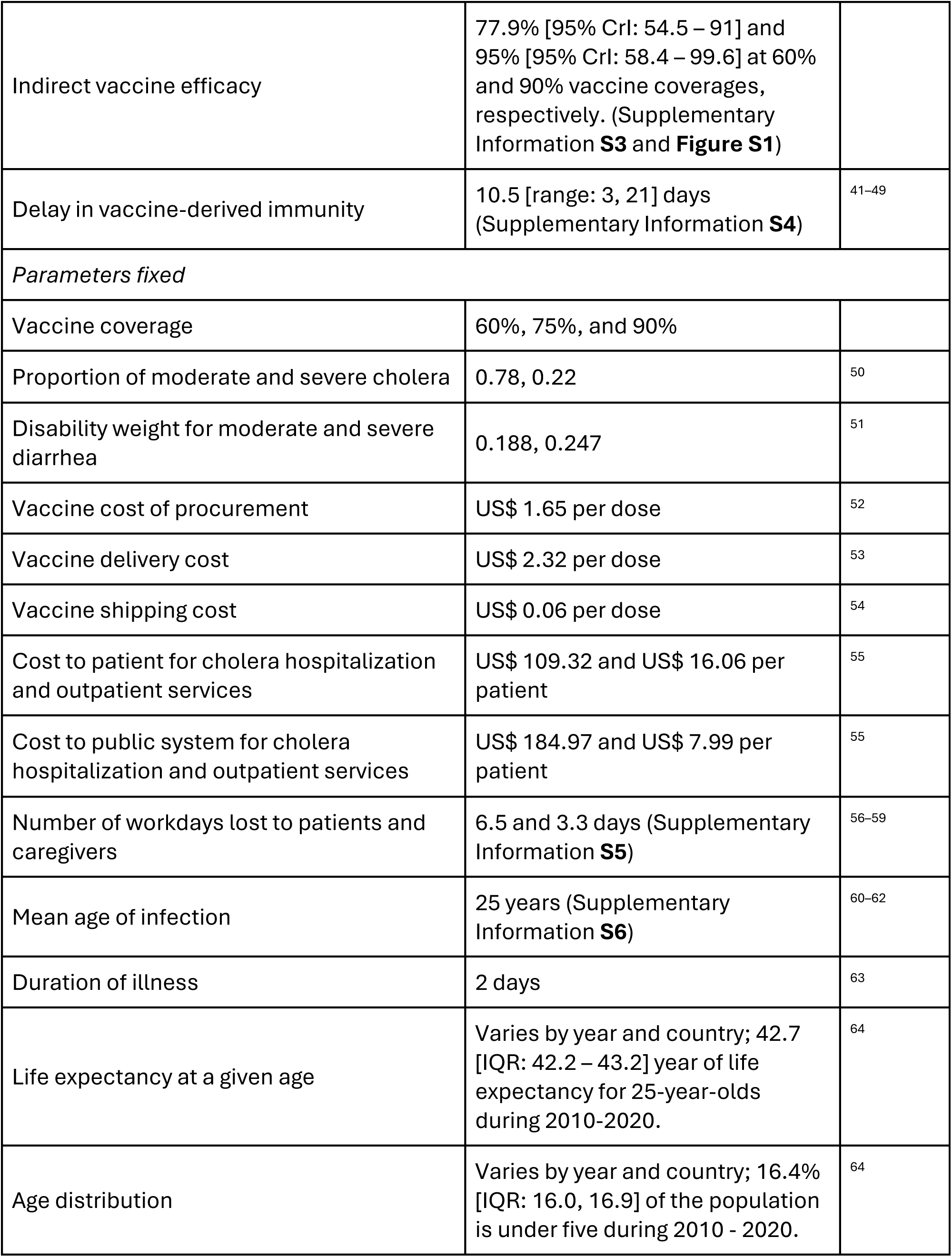
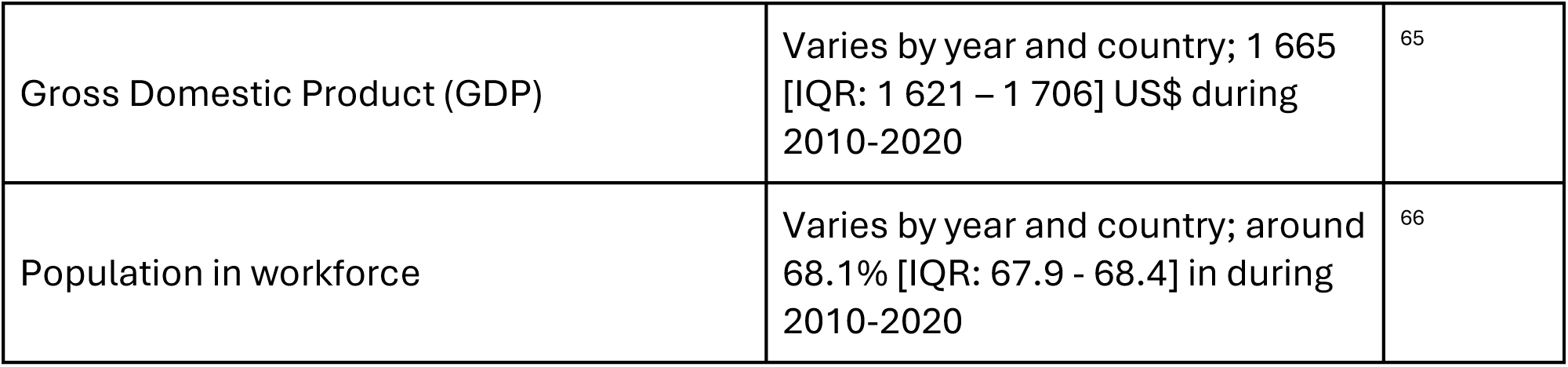
Model parameters.

**Table 2.** Model settings for ORI (single-dose OCV campaign) settings.

|  |  |
| --- | --- |
| OCV campaign feature | Model settings that were tested |
| Campaign coverage | Target population included the entire population of the affected administrative unit. Three coverage levels were modeled: 60%, 75%, and 90%. |
| Campaign triggers | (i) Outbreak onset–triggered: ORI initiated in 1–15 weeks after outbreak onset, in 1-week increments; implementation set at the midpoint of the specified week.<br>(ii) Case-triggered: ORI initiated once cumulative cases reached a predefined threshold (1, 5, 10, 30, 50, 70, 90, 140, 190, 240, 290 cases), followed by an additional 1–15 week implementation delay. |
| Vaccination rounds | Single-round vaccination campaign per outbreak |
| Outbreak data | Analyses performed at both the individual-outbreak level and in aggregated subsets selected by outbreak characteristics, including attack rate, duration, size, and case-fatality ratio. |

**Table 3.**
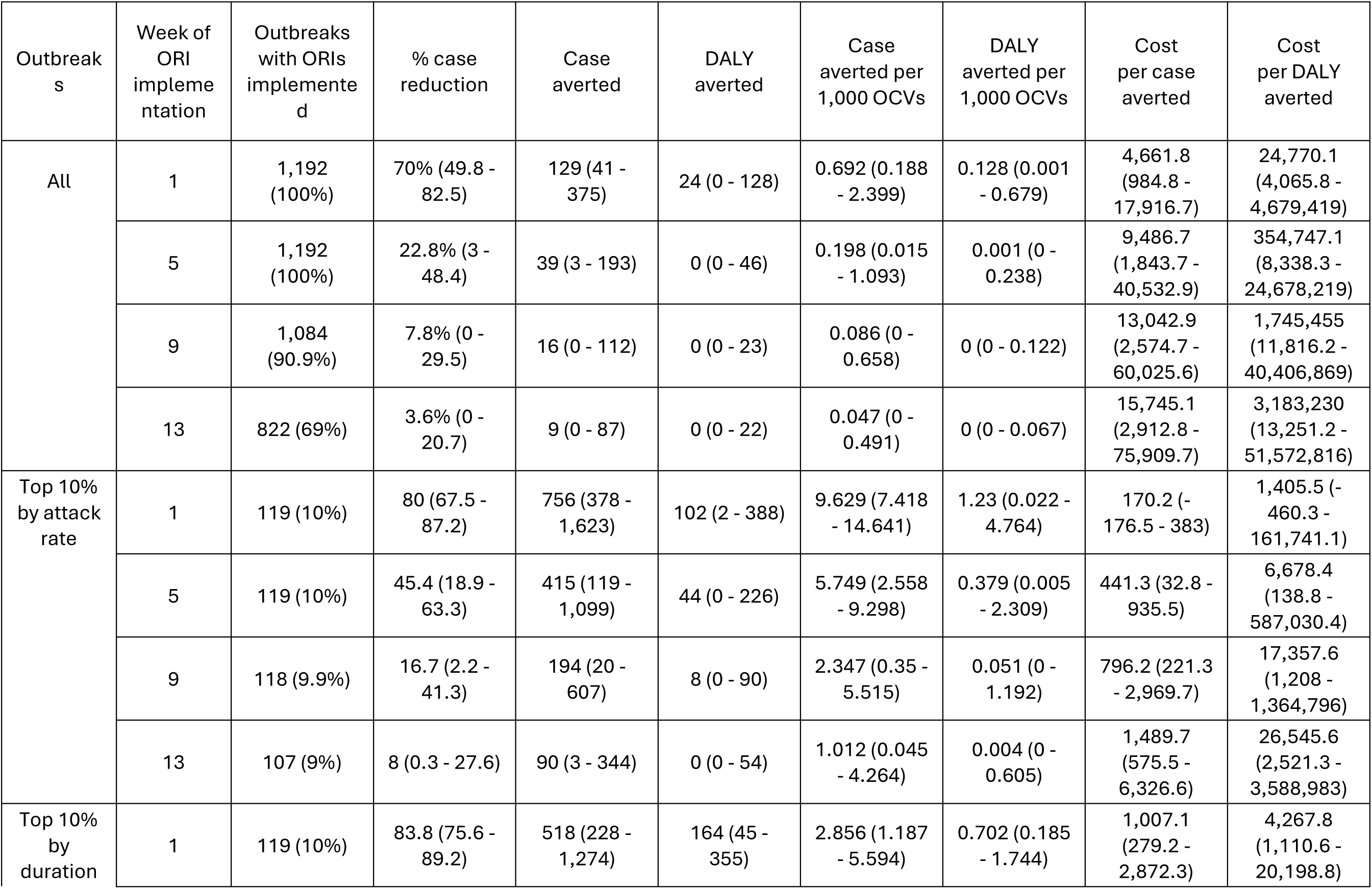
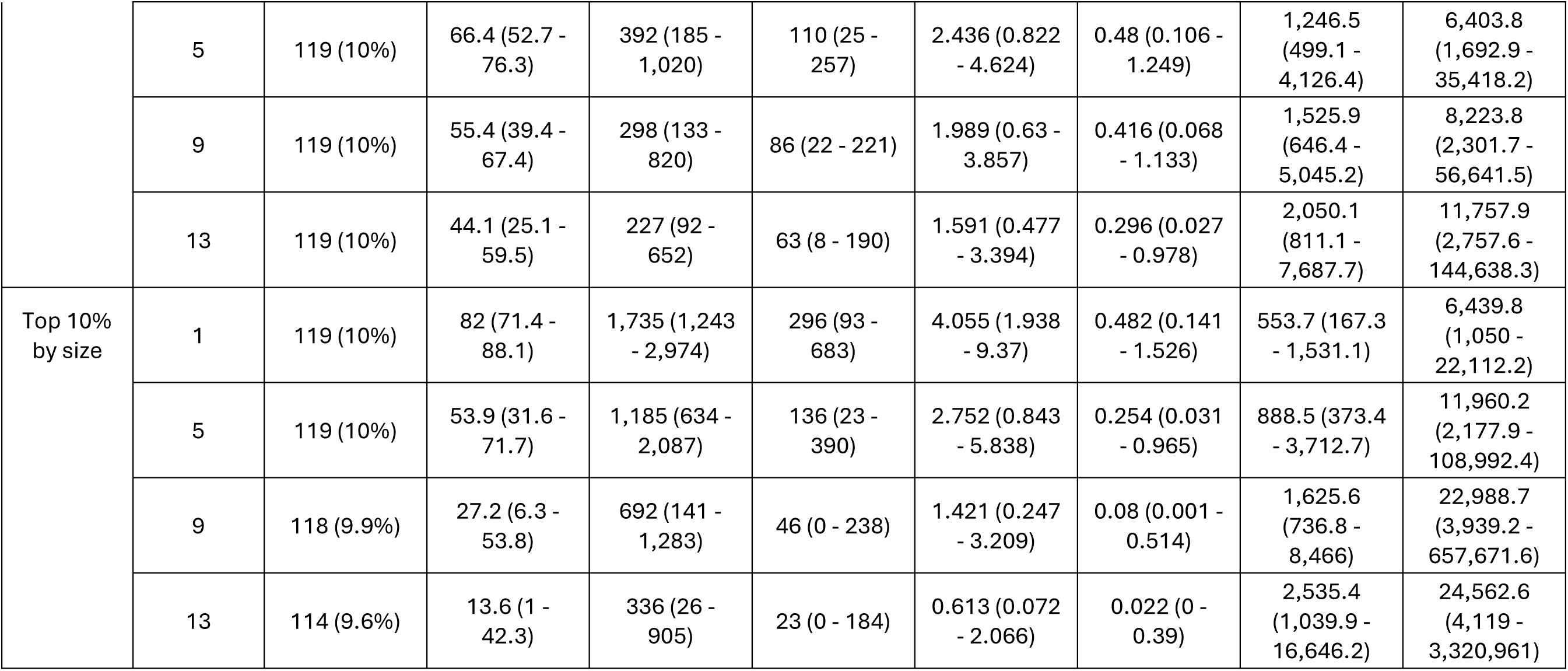
Impact of ORI with 75% vaccine coverage under the outbreak onset-triggered scenario. Values represent medians with interquartile ranges (IQRs).

To estimate the global impact of the ORI, we aggregated results across outbreaks assuming a constrained OCV supply of approximately 100,000 to 200 million doses, roughly reflecting the scale of global stockpile deployments.^36^ Because this supply was insufficient to vaccinate all outbreak populations, we modeled allocation strategies across all or selected outbreaks prioritized by characteristics such as attack rate, duration, size, or case-fatality ratio. Outbreaks were sampled without replacement and assigned enough vaccine to reach their target coverage. Sampling continued until the cumulative number of doses matched the supply. This allocation was repeated 200 iterations.

The model captures on the immediate, within-outbreak impact of ORI and does not account for potential long-term benefits, such as preventing or mitigating future outbreaks. This assumption reflects the temporary population-level protection expected from single-dose campaigns, given waning immunity and population mobility.^9^ Moreover, we modeled each population independently, without cross-population transmission or vaccination spillover.

### Campaign triggers and assumptions

ORIs were triggered at the outbreak onset (outbreak onset-triggered) or when the cumulative number of reported cases exceeded a predefined threshold (case-triggered) (**Table 2**). The outbreak onset-triggered approach isolates the effect of response delay, clarifying how timing influences ORI impact. The case-triggered approach reflects potentially more realistic conditions in which uncertainty in surveillance or confirmation delays may postpone the start of campaigns. Under this scenario, identical case thresholds may trigger earlier responses in rapidly growing outbreaks and later responses in slower ones.

Vaccine coverage was varied across 60 %, 75 %, and 90 % of the affected administrative unit’s population, consistent with ranges observed in global OCV campaigns (2013-2018)^67^ and a recent campaign in Ethiopia.^68^ We selected 75% as the representative single-dose coverage, reflecting levels typically achieved in humanitarian crises or outbreaks (Supplementary Information **S7**). The delay between vaccination and the onset of protection was based on observed seroconversion times (proxy for immune response),^41–49^ ranging from 3 to 28 days (most often 7 to 14 days) after the first dose of a two-dose regimen^41,43–46^ (Supplementary Information **S4**).

### Cost-effectiveness analysis

Cost-effectiveness was assessed as the cost per case, death, and DALY averted. Cost estimates were derived from published literature (**Table 1**; Supplementary Material **S8**). We adopted a societal perspective on cost, incorporating both direct costs (e.g., vaccine procurement and healthcare utilization) and indirect costs (e.g., productivity losses). All costs were converted to 2023 US dollars, adjusting for inflation using annual consumer price index changes for sub-Saharan Africa through 2023.^69–71^

## Results

### Outbreak data

We identified 1,192 cholera outbreaks across sub-Saharan Africa between 2010 and 2020. Cholera incidence displayed marked spatial and temporal heterogeneity across Africa.

Monthly time series indicated that most outbreaks occurred in Eastern, Western, and Central Africa (**Figure 2A**). Outbreaks during 2010–2015 were concentrated in Western Africa, whereas those during 2016–2021 were more frequent in Eastern Africa. Central Africa experienced outbreaks more consistently throughout the period. Spatial heterogeneity was also evident at finer geographic scales (**Figure 2B**). Outbreaks typically peaked around week 5 (mean peak at week 6, median week 4), and approximately 88% peaked before week 10 (**Figure 2C**). Outbreak durations ranged from 5 to 93 weeks, with sizes from 3 to 14,522 suspected cases and the attack rates from 0·005 to 112 rates per 10,000 populations (**Figures 2D-E**). Among the 152 outbreaks reporting at least one laboratory-confirmed case, duration ranged from 5 to 56 weeks, outbreak sizes from 10 to 7,726 suspected cases, and attack rates from 0.005 to 19 cases per 10,000 population. The outbreaks we excluded showed similar patterns and were simply larger on average, suggesting our analyzed dataset is broadly representative of cholera epidemiology in the region (**Figure S2**).

**Figure 2.**
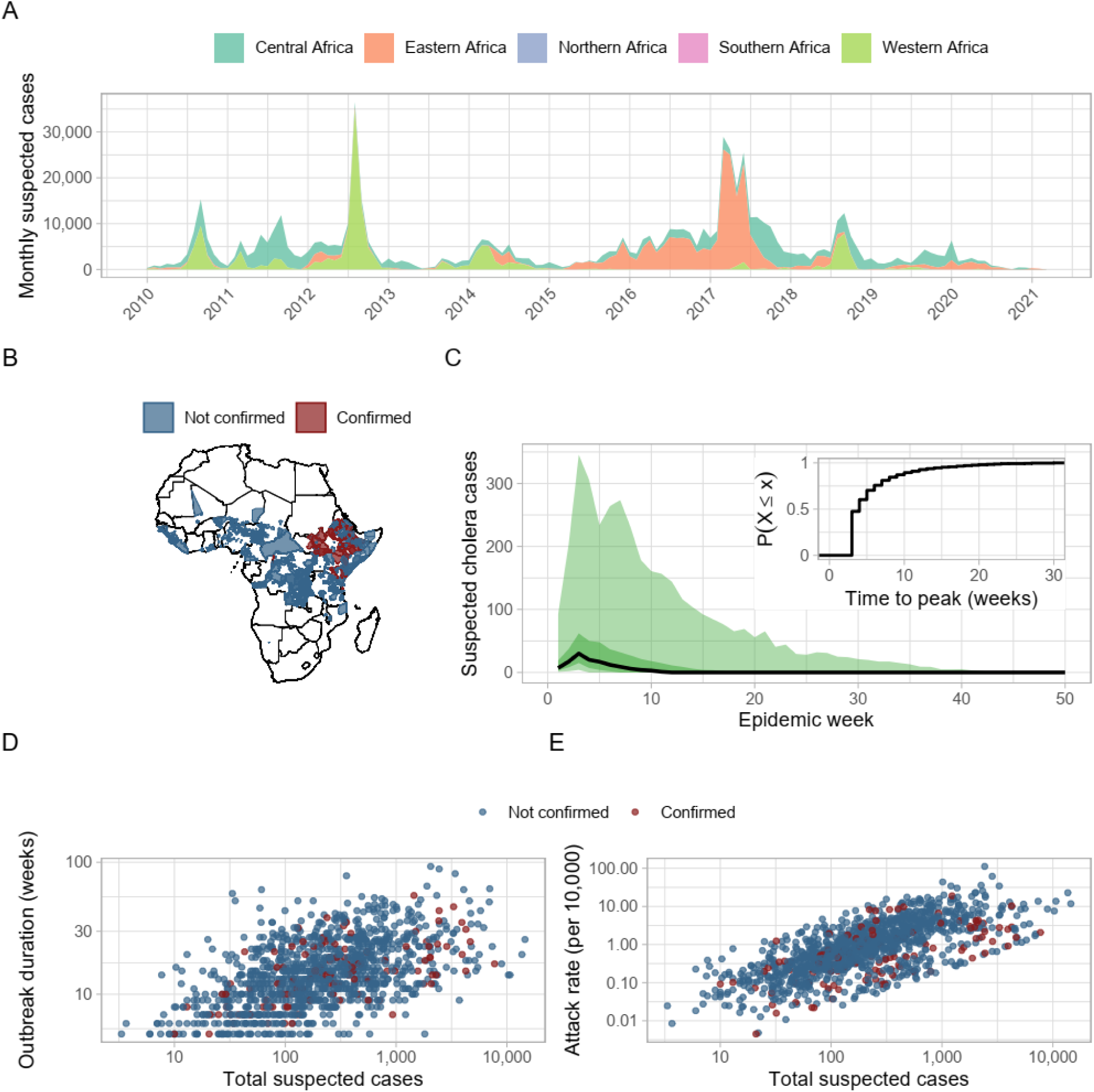
Characteristics of cholera outbreaks in sub-Saharan Africa, 2010–2020. Outbreak data set. (A) Monthly time series of suspected cholera cases, stacked by African subregion. (B) Geographic distribution of outbreaks by administrative levels 1, 2, and 3. Darker shading indicates areas with repeated outbreaks. (C) Median epidemic curve with 95% central percentiles; inset shows the cumulative probability distribution of time to peak (in weeks). (D) Total number of suspected cases versus outbreak duration, both on a logarithmic scale. (E) Total number of suspected cases versus attack rate (per 10,000 population), both on a logarithmic scale.

### ORIs in individual outbreaks

#### Impact across campaign timing and coverage

In the outbreak-onset-triggered scenario (i.e. vaccination triggered at the start of an outbreak), an ORI launched in week 1 at 75% coverage averted a median of 70% [IQR: 49.8– 82.5] of cases—corresponding to 129 cases [IQR: 41–375] averted (**Figure 3A**). Higher coverage yielded greater benefit: at week 1, the median cases averted were 120, 129, and 135 for 60%, 75%, and 90% coverages, respectively. By week 9, median cases averted fell dramatically to 15, 16, and 17 cases (under 9% of total cases at any coverage level).

**Figure. 3.**
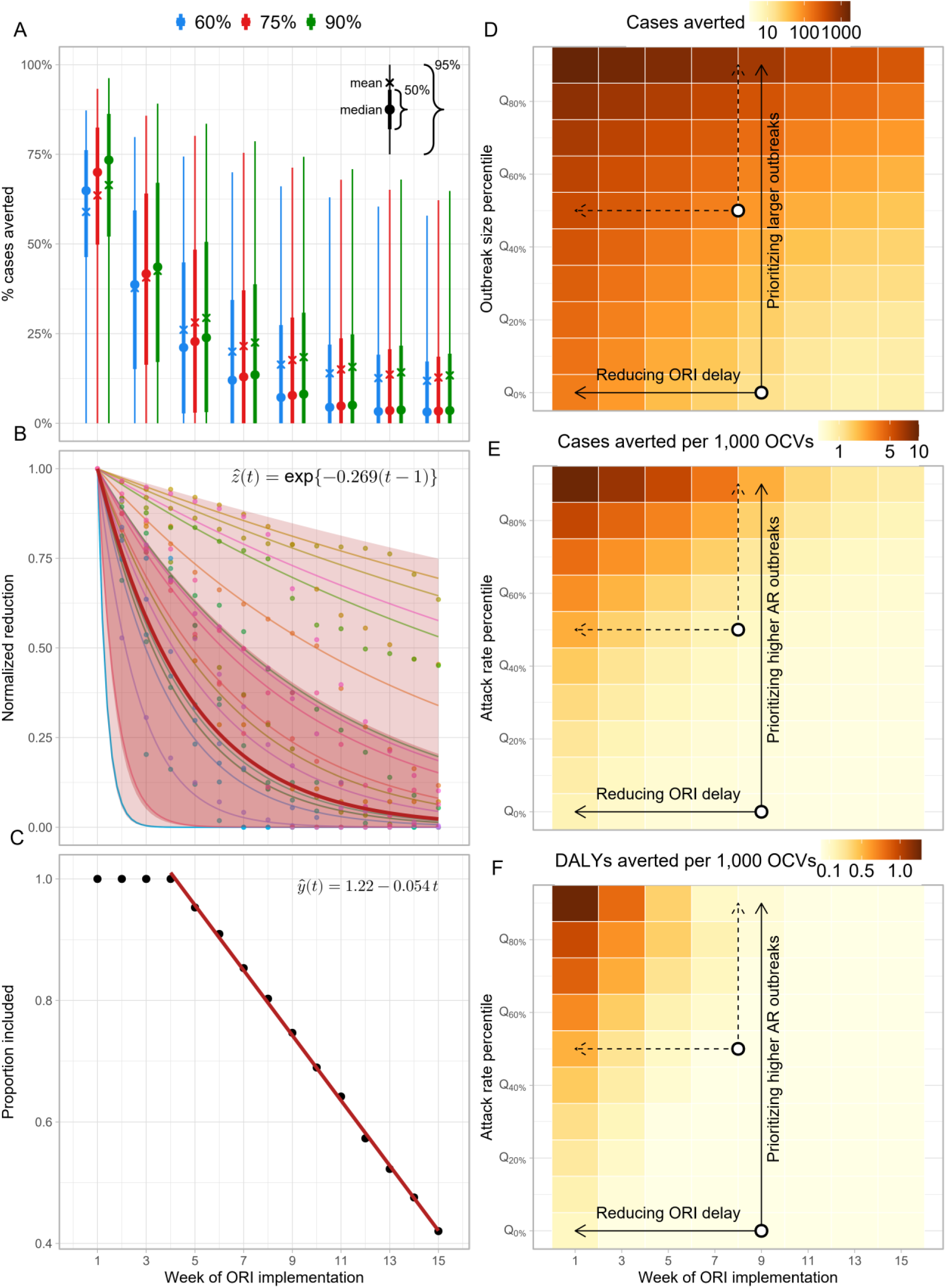
Impact of outbreak response immunization (ORI). (A) Percentage cases averted for ORIs implemented in week 1 to week 15 under vaccine coverage of 60%, 75%, and 90%. (B) Exponential decay fits to the proportion of cases averted, normalized to the week-1 value, *ẑ*(*t*), under 75% vaccine coverage: thin colored curves show 30 representative outbreaks; the thick red curve is the median; darker shading denotes the interquartile (50%) envelope and lighter shading the 95% envelope. (C) Proportion of outbreaks, *ŷ*(*t*), eligible for analysis at each week (i.e., outbreaks long enough for an ORI to have occurred by that week). (D-F) Median cases averted per 1, 000 OCVs across delay in ORI implementation under 75% vaccine coverage for subsets defined by (D) attack rate, (E) outbreak duration, and (F) outbreak size. Y-axis labels of the form, Q_X%_, denote subsets containing outbreaks at or above the X*th* percentile of the corresponding characteristic.

Substantial variability in outcomes across outbreaks remained for each delay, reflecting heterogeneity in outbreak characteristics and campaign parameters. Despite variability, the relationship between delay and impact followed a clear exponential decay (*R*^2^= 0.90 [IQR: 0.78–0.96]) (**Figure 3B**). Among outbreaks lasting >=7 weeks (91% of outbreaks), for which ORIs could be implemented as late as week 9, the decay rate was 0.27 [IQR: 0.11–1.49] per week, corresponding to a weekly decline of 23.6% [IQR: 10.7–77.4] % and a halving time of 2.6 [IQR: 0.5–6.1] weeks. The decay was slower in longer outbreaks (weekly decline of 19.2 [IQR: 9–61.7] for outbreaks lasting >= 17 weeks; 32% of outbreaks). Delay also caused some outbreaks to end because vaccination began (**Figure 3C**). From week 5 onward, an additional 5.4% of outbreaks were missed per week of delay, reaching around 25.3% by week 9.

### Efficiency and cost effectiveness across campaign timing and prioritization

Prioritizing larger outbreaks produced the greatest absolute case reduction (**Figure 3D; Figure S3** in the Supplementary Information) whereas prioritizing high-attack-rate outbreaks maximized ORI efficiency (**Figures 3E–F**). Targeting the top 10% of outbreaks by size increased the median number of cases averted to 682 cases (vs. 16 under random targeting) for week-9 ORIs, and to 1,725 cases for week-1 ORIs. Prioritizing the top 10% by attack rate increased the ORI efficiency approximately to 9.58 cases (1.28 DALYs) and 2.39 (or 0.06 DALYs) averted per 1,000 doses for week-1 and week-9 ORIs, respectively— equivalent to 14–28-fold (10–397-fold for DALYs) increase compared to estimates based on all outbreaks.

Prioritizing by size or duration also improved efficiency but to a lesser extent (**Figure S3**). Percentage of cases averted, which was not normalized by doses, was most sensitive to outbreak duration. The median percentage of cases averted were 83.8%, 82%, and 80.1% for outbreaks in the top decile of duration, size, and attack rate, respectively, for week-1 ORIs, declining to 55.2%, 26.7%, and 17.1% by week 9.

**Table 3** summarizes the impact, efficiency, and cost effectiveness of the outbreak onset-triggered ORI with 75% vaccine coverage (results for other coverage levels and delays appear in **Table S1**). As delays increased, the impact and the efficiency of ORIs declined, while cost-effectiveness improved. The cost per case averted increased from 4,662 [IQR: 985 - 17,917] for week-1 ORIs to US$13,043 [IQR: 2,575–60,026] for week-9 ORIs.

Corresponding costs per DALY averted rose from US$24,770 [IQR: 4,066–4,679,419] to US$1,745,455 [IQR: 11,816–40,406,869], exceeding the median GDP per capita in sub-Saharan Africa ($1,665^65^) in 2010 - 2020. Impact, efficiency, and cost effectiveness were consistently greater among outbreaks with higher attack rates, longer duration, or larger size. In the top 10% by duration, percentage of case averted reached 83.8% [IQR: 75.6–89.2] and 55.4% [IQR: 39.4–67.4] for week-1 and week-9 ORIs, respectively. Targeting the top quartile by attack rate yielded the most favorable efficiency and cost effectiveness outcomes. Week-1 and week-9 ORIs cost US$ 1,406 [IQR: -460–161,741] and US$ 17,356 [IQR: 1,208–1,364,796] per DALY averted, respectively— equivalent to 0.8 and 10.4 times the regional GDP per capita, and representing 94% and 99% reductions relative to estimates based on all outbreaks.

### Case-triggered scenario

For the case-triggered scenario (vaccination triggered only after a threshold number of cases), the impact, efficiency, and cost effectiveness declined with increasing implementation delays and higher case thresholds triggering vaccination (**Table 4**). Median percentage of cases averted for week-1 ORIs ranged from 70% (comparable to the outbreak-onset-triggered scenario, to below 35% in case ∼140 cases were required to trigger a response. An exponential decay model (*R*^2^ = 0.87 [IQR: 0.77–0.93]) predicted that case averted decreased by 6.4% [IQR: 2.5–14.2] for every 10 additional cases missed before response initiation, corresponding to halving every ∼105 additional cases [IQR: 45.3–272.7].

**Table 4.**
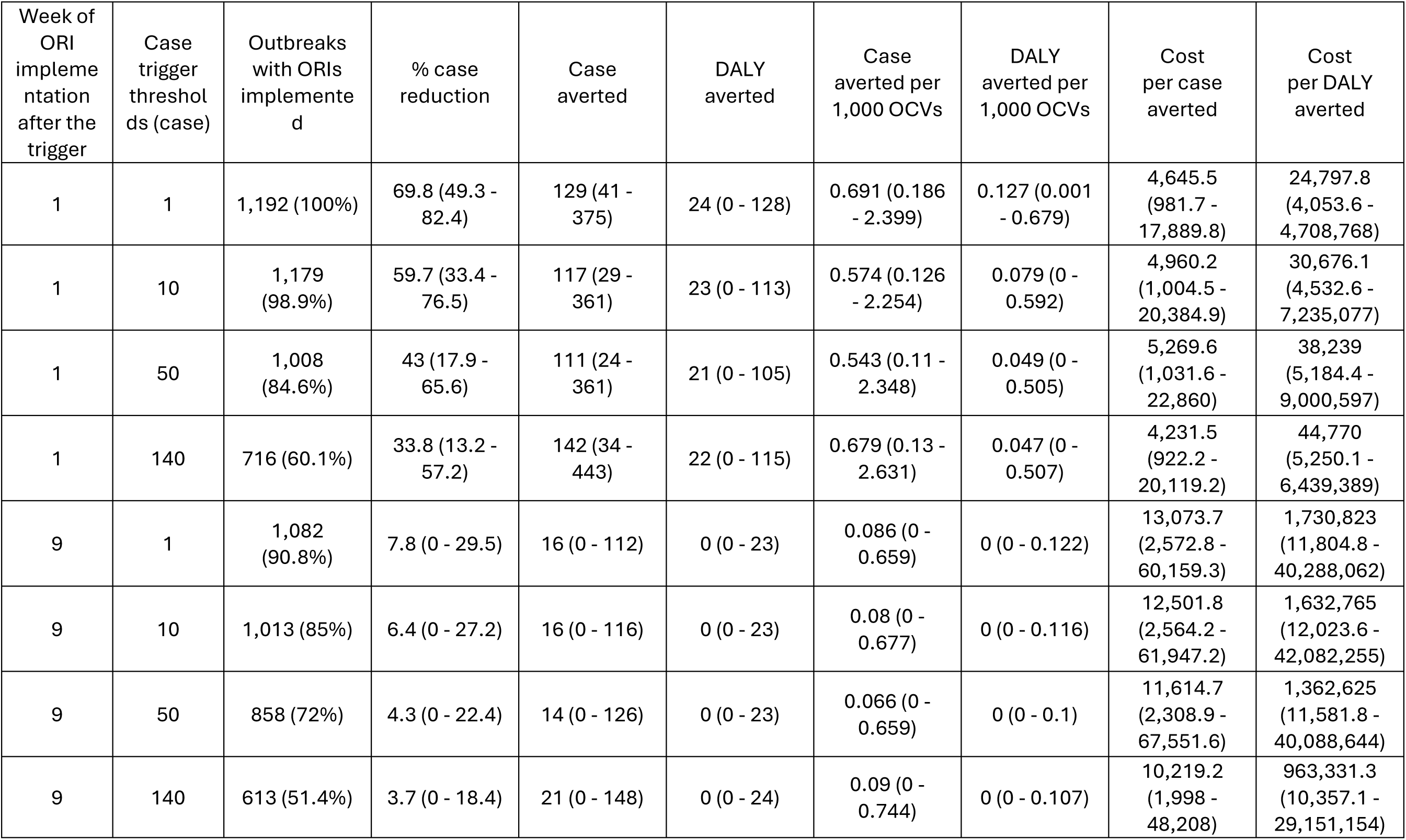
Impact of ORI with 75% vaccine coverage under the case-triggered scenario. Values represent medians with interquartile ranges (IQRs).

| Week of ORI implementation after the trigger | Case trigger thresholds (case) | Outbreaks with ORIs implemented | % case reduction | Case averted | DALY averted | Case averted per 1,000 OCVs | DALY averted per 1,000 OCVs | Cost per case averted | Cost per DALY averted |
| --- | --- | --- | --- | --- | --- | --- | --- | --- | --- |
| 1 | 1 | 1,192 (100%) | 69.8 (49.3 - 82.4) | 129 (41 - 375) | 24 (0 - 128) | 0.691 (0.186 - 2.399) | 0.127 (0.001 - 0.679) | 4,645.5 (981.7 - 17,889.8) | 24,797.8 (4,053.6 - 4,708,768) |
| 1 | 10 | 1,179 (98.9%) | 59.7 (33.4 - 76.5) | 117 (29 - 361) | 23 (0 - 113) | 0.574 (0.126 - 2.254) | 0.079 (0 - 0.592) | 4,960.2 (1,004.5 - 20,384.9) | 30,676.1 (4,532.6 - 7,235,077) |
| 1 | 50 | 1,008 (84.6%) | 43 (17.9 - 65.6) | 111 (24 - 361) | 21 (0 - 105) | 0.543 (0.11 - 2.348) | 0.049 (0 - 0.505) | 5,269.6 (1,031.6 - 22,860) | 38,239 (5,184.4 - 9,000,597) |
| 1 | 140 | 716 (60.1%) | 33.8 (13.2 - 57.2) | 142 (34 - 443) | 22 (0 - 115) | 0.679 (0.13 - 2.631) | 0.047 (0 - 0.507) | 4,231.5 (922.2 - 20,119.2) | 44,770 (5,250.1 - 6,439,389) |
| 9 | 1 | 1,082 (90.8%) | 7.8 (0 - 29.5) | 16 (0 - 112) | 0 (0 - 23) | 0.086 (0 - 0.659) | 0 (0 - 0.122) | 13,073.7 (2,572.8 - 60,159.3) | 1,730,823 (11,804.8 - 40,288,062) |
| 9 | 10 | 1,013 (85%) | 6.4 (0 - 27.2) | 16 (0 - 116) | 0 (0 - 23) | 0.08 (0 - 0.677) | 0 (0 - 0.116) | 12,501.8 (2,564.2 - 61,947.2) | 1,632,765 (12,023.6 - 42,082,255) |
| 9 | 50 | 858 (72%) | 4.3 (0 - 22.4) | 14 (0 - 126) | 0 (0 - 23) | 0.066 (0 - 0.659) | 0 (0 - 0.1) | 11,614.7 (2,308.9 - 67,551.6) | 1,362,625 (11,581.8 - 40,088,644) |
| 9 | 140 | 613 (51.4%) | 3.7 (0 - 18.4) | 21 (0 - 148) | 0 (0 - 24) | 0.09 (0 - 0.744) | 0 (0 - 0.107) | 10,219.2 (1,998 - 48,208) | 963,331.3 (10,357.1 - 29,151,154) |

### ORIs across multiple outbreaks

#### Impact across campaign timing, OCV doses, and prioritization

Assuming 100,000 to 200 million OCV doses, the number of outbreaks eligible for ORI ranged from 1 to 559 when random targeting. Because outbreaks varied in population size, prioritizing larger outbreaks reduced the number targeted (435 [IQR: 420-453] outbreaks for 200 million doses), whereas prioritizing those with higher attack rate increased it (776 [IQR: 769–785]), reflecting the tendency of high-attack-rate outbreaks to be smaller (**Figure 4A**). Total cases averted increased with both the number of doses deployed and prioritization (**Figure 4B**). For ORIs implemented in week 9, the median number of cases averted per 200 million doses was 72,362 [IQR: 66,407–79,326] under random targeting, compared with 89,347 [IQR: 83,640 - 95,170], 96,929 [IQR: 91,991–104,011], and 156,732 [IQR: 154,407–158,981] when prioritizing by outbreak size, duration, and attack rate, respectively. When the delay was shortened to 1 week, the corresponding medians increased to 200,283, 247,062, 260,528, and 422,804, respectively.

**Figure 4.**
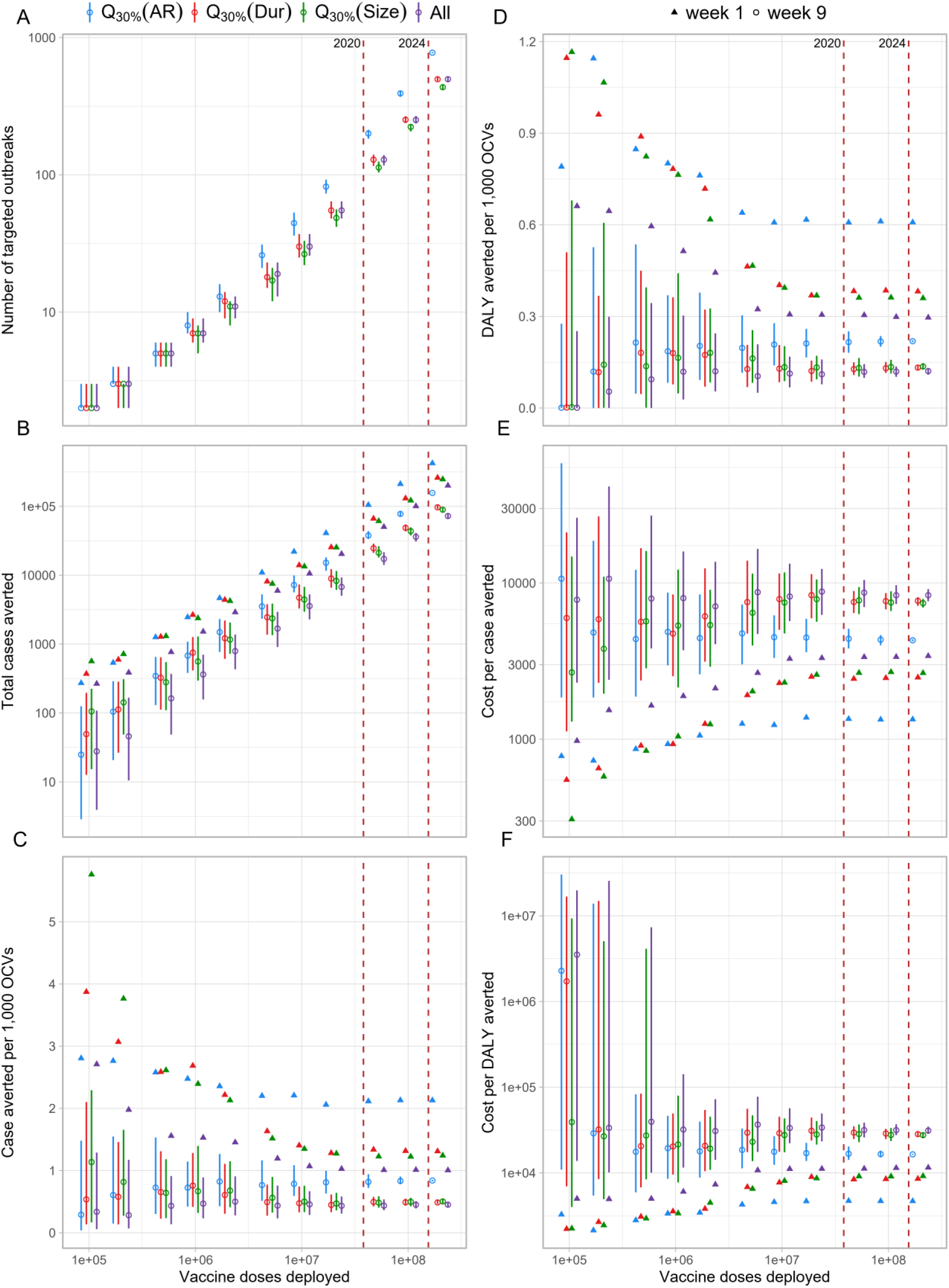
Impact, efficiency, and cost effectiveness of ORIs under 75% coverage for subsets of outbreaks. (A) The number of outbreaks targeted against the number of OCV doses deployed. The red dashed lines indicate the number of OCV doses deployed for reactive vaccination up to 2020 (maximum year in our dataset) and 2024. (B) The number of cases averted. (C) The number of cases averted per 1,000 OCVs. (D) DALY averted per 1,000 OCVs. (E) Cost per case averted. (F) Cost per DALY averted. Empty circles and filled upward triangles represent median values across 200 samples of outbreaks for ORIs implemented in week 1 and week 9, respectively. Error bars show interquartile ranges for the outcomes from ORIs implemented in week 9. Q[*x*%] (*x*), denote subsets containing outbreaks at or above the *x*th percentile of the variable *x*.

### Efficiency and cost effectiveness across campaign timing, OCV doses, and prioritization

ORI efficiency varied widely at low vaccine supply (<= 200,000 doses) but converged as supply increased (**Figure 4C**). Targeting high-attack-rate outbreaks consistently achieved the greatest efficiency, exceeding 2 cases averted per 1,000 doses for week-1 ORIs and a bit lower than 1 case per 1,000 doses for week-9 ORIs, while other targeting strategies remained lower. The pattern for DALYs averted per 1,000 doses mirrored that of cases averted per 1,000 doses, though with smaller magnitude—approximately 0.6 for high-attack-rate targeting and 0.3–0.4 for other targeting strategies for week 1 ORIs (**Figure 4D**).

Cost effectiveness improved both with both shorter delays and targeted deployment (**Figures 4E-F**). The cost per case averted decreased from US$ 8,352 [IQR: 7,641–9,140] under random targeting to US$ 4,298 [IQR: 4,237–4,368] when high-attack-rate outbreaks were prioritized with week-9 ORIs, and further to US$ 3,410 [IQR: 3,173–3,615] and US$ 1,340 [IQR: 1,321–1,362] for random and high-attack-rate targeting, respectively, with week-1 ORIs (**Figure 4E**). Similarly, cost per DALY averted declined from US$ 31,157 [IQR: 28,533-34,696] to US$ 16,465 [IQR: 16,205–16,752] for week-9 ORIs, and from US$ 11,531 [IQR: 10,919–12,479] to US$ 4,688 [IQR: 4,624–4,786] for week-1 ORIs (**Figure 4F**).

## Discussion

Our study showed the substantial potential of OCVs to reduce the burden of cholera outbreaks, based on modeling of 1,192 outbreaks in sub-Saharan Africa from 2010 to 2020. Timing of ORI was critical: median cases averted declined from 70% with week-1 implementation, halving every 2.6 weeks of delay. We also found that prioritizing outbreaks—especially those with high attack rates—substantially improved ORI efficiency and cost-effectiveness. Finally, while delayed ORIs may appear to have limited impact at the level of individual outbreaks, their aggregate effect can be meaningful when larger, longer, or higher-burden outbreaks are included.

The benefit of ORI varied widely across weeks of delay, with percentage of cases averted often ranging from near 0% to 100%, and with substantial overlap between different delay periods (see **Figure 3A**). This variability reflects not only differences in campaign parameters (coverage and delay) but also diverse outbreak characteristics, as shown in our stratified analyses by attack rate, duration, and size. These findings suggest that losses from delayed implementation can be partly mitigated by prioritizing outbreaks that are larger, longer lasting, or have higher attack rates. Consistent with previous analyses highlighting the benefits of selective targeting in sub-Saharan Africa,^5,72^ these results underscore the value of strategic prioritization. Allocating limited OCV supplies to outbreaks most likely to become extensive or prolonged could markedly increase the impact, efficiency, and cost effectiveness of ORIs.

Although such prioritization assumes an ability to predict outbreak trajectory at the time of response, this may be increasingly feasible using environmental and socioeconomic indicators, and WASH conditions, and historical outbreak trends.^21,73–76^ As predictive models and surveillance systems improve, they can guide more targeted and efficient reactive vaccination.

Our analysis of case-triggered ORIs further underscores the importance of early detection as well as rapid implementation. For ORIs initiated in week 1 of an outbreak (week 1 ORIs), the median percentage of cases averted declined from 70% when vaccination was triggered at outbreak onset to below 35% when triggered only after more than 105 cases had already occurred—equivalent to surveillance missing the first 105 cases or so and delaying the response well beyond the true first week. These findings highlight the need for robust surveillance systems capable of rapidly detecting and confirming cholera cases, alongside operational and logistical capacity to deliver vaccines and initiate campaigns immediately once response thresholds are reached.

At the aggregate level, the overall impact of ORI did not scale linearly with the median outbreak-level estimates. When successive ORIs were implemented across multiple outbreaks, large and long-duration outbreaks—which yielded greater benefits from ORIs— contributed disproportionately to the overall benefit, moving the overall median upward.

Assuming 200 million OCV doses (comparable to the total deployed globally since 2013), our model estimated that random targeting would avert around 0.45 cases per 1,000 doses (compared to 0.1 at the outbreak level), increasing to 2.1 when prioritizing high-attack-rate outbreaks, equivalent to about 422,000 cases averted for week-1 ORIs.

Our estimates of impact, efficiency, and cost-effectiveness are broadly consistent with, but likely more representative than, previous studies that focused on a few large-scale outbreaks.^2,3,5,7,8,10,12,15,19,77^. For instance, analyses based on a few outbreaks have reported roughly 30–90% case reductions from reactive campaigns in Bangladesh, Zimbabwe, Guinea, Haiti, Chad, and Thailand,^1,5,13,14^ often assuming high coverage or early response. By incorporating over a thousand outbreaks of varying epidemiologic characteristics, our approach provides regionally generalizable estimates across realistic operational settings. The ORI efficiency in our study (2.1 cases averted per 1,000 doses when prioritizing high- attack-rate outbreaks) aligns with the estimates from Xu et al.,^78^ who reported 2.6–11 cases averted per 1,000 fully vaccinated persons (two doses) in preventive campaigns across sub-Saharan Africa. Although higher efficiencies have been reported from campaigns in Bangladesh,^79^, Indonesia, Nigeria, and Uganda^80^, and Guinea, Haiti, Zimbabwe, Haiti,^14^ these likely reflect conditions not representative of most outbreaks. Likewise, our estimated cost per DALY averted (∼US $ 24,770 and US $ 2,977 for random targeting and the top quartile by attack rate with week-1 ORIs) falls within previously reported ranges (US $1,800–21,000 per DALY averted) for two-dose preventive campaigns in sub-Saharan Africa.^72^

This study has several limitations. First, we assumed uniform vaccine coverage within administrative units, which may underestimate the efficiency of spatially focused strategies such as case-area targeted interventions^5,81–83^ or district-level geographic targeting.^72,78^ Nonetheless, our findings showing higher efficiency of ORIs in outbreaks with high attack rates suggest that these conditions can approximate the benefits of spatially targeted approaches. Conversely, unaccounted vaccine wastage^84^ could lead to overestimation of ORI efficiency and cost effectiveness. Second, our parameterization of indirect protection relied on data from Bangladesh,^1,39^ which may not fully generalize to sub-Saharan African contexts; nevertheless, the consistency between our estimates and the outcomes from the dynamic model (Supplementary Information **S9; Figures S5-S7**) suggests that modeled indirect effects are robust. Third, we modeled each outbreak independently and thus the impact of ORI was confined to the specific outbreak in which the ORI was implemented. In reality, ORIs might reduce future outbreaks in the same or neighboring areas given that two-dose OCVs provide protection for up to 4-5 years.^85–88^ Due to a lack of geographic details for each outbreak and epidemiologic links between the outbreaks, we could not evaluate the downstream or spatial spillover effects. Fourth, as roughly half of reported suspected cholera cases are not true *Vibrio cholerae* infections.^89^ our reliance on suspected case data may overestimate impact; however, underreporting of cholera in sub-Saharan Africa may offset this bias^90^. Fifth, we assumed ORIs would not be implemented if more than two weeks had elapsed after outbreak resolution, whereas in practice, reactive campaigns may still occur. Sensitivity analyses confirmed a monotonic decline in ORI impact and efficiency as the delay to stop ORI after outbreak end increased (**Table S2**). Sixth, our outbreak dataset was generated using standardized but operational definitions that vary by context. Alternative outbreak criteria could yield different datasets and, consequently, different estimates. The main analysis excluded outbreaks from administrative units with populations that were outside of typical range of OCV deployments (16,500–5,000,000). To explore a broader range of outbreak settings, we relaxed these criteria and included outbreaks occurring in populations between 5,000 and 10,000,000. Under this more inclusive scenario, the median and interquartile ranges remained largely unchanged (**Table S3**). Finally, our simulation of OCV deployment from the global stockpile simplified several realistic details—such as characteristics of outbreaks selected, variability in coverage, and the full distribution of implementation delays—to highlight the disproportionate impact of long and large outbreaks to overall ORI impact and the benefits of prioritization by outbreak characteristics. These results should therefore be interpreted qualitatively.

## Conclusion

Our study quantifies the health and economic benefits of timely and strategically targeted OCV outbreak response across diverse epidemiological settings in sub-Saharan Africa.

Rapid ORI implementation and prioritization of large, prolonged, or high–attack-rate outbreaks can maximize health gains and cost-effectiveness, even under vaccine supply constraints. Strengthening surveillance, improving predictive tools, and ensuring rapid vaccine deployment will be essential to realize the full potential of reactive cholera vaccination.

## Data Availability

All data produced are available online at: https://github.com/kimfinale/cholera-outbreak-vaccine-impact-modeling

## Acknowledgement

This work was supported, in whole or in part, by the Bill & Melinda Gates Foundation [INV-009125]. The conclusions and opinions expressed in this work are those of the authors alone and shall not be attributed to the Foundation. Under the grant conditions of the Foundation, a Creative Commons Attribution 4.0 License has already been assigned to the Author Accepted Manuscript version that might arise from this submission. Please note works submitted as a preprint have not undergone a peer review process.

## Author contributions

J-HK and ECL conceptualized the study. J-HK led the study design and analysis. J-HK and MD performed the statistical analysis and drafted the initial manuscript. ECL and QZ collected and prepared the data for analysis. ECL, ASA, and JHK provided critical input on methodology. MD conducted the literature review. All authors participated in the interpretation of the findings, critically reviewed the manuscript for intellectual content, and approved the final version for submission.

## S1. Vaccine impact

Following the notations used in the previous study,^1^ we define population *A*, in which a fraction, *π*, of the population is vaccinated and the rest, 1 − *π*, of the population remains unvaccinated. We define another population, *B*, in which no one was vaccinated. Let *C*_*A*0_ and *C*_*A*1_ denote cumulative incidence in the unvaccinated and vaccinated individuals of population *A*. Similarly, let *C*_*B*0_denote cumulative incidence in population *B*. Four measures of vaccine effectiveness—direct (DVE), indirect (IVE), total (TVE), and overall (OVE)—can be defined as follows:

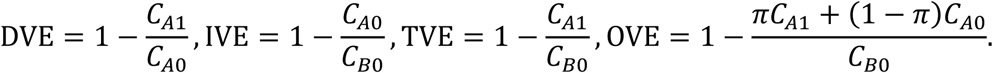

We see that the following relationships hold true.

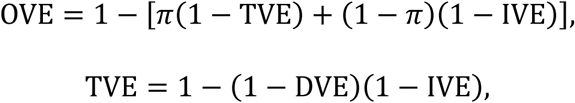

The impact of vaccine was modeled based on the definition of overall effectiveness, given the direct and indirect vaccine effectiveness, of which estimates are available. The cumulative incidence, 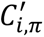, for age group *i* in a population in which a proportion, *π*, of population was vaccinated can be defined as follows:

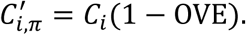

where *C*_*i*_, represents the incidence in the population without vaccines. We replace OVE in the equation using the DVE and IVE, for which estimates are available, based on the relationship we stated above:

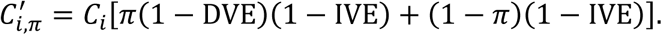

## S2. Indirect vaccine effectiveness

Indirect vaccine effectiveness (IVE) of the oral cholera vaccine (OCV) was estimated using incidence rates among unvaccinated individuals residing in clusters with varying vaccine coverage levels, as observed in a randomized trial conducted in Matlab, Bangladesh.^2^

Vaccine coverage rates were reported as a range and the mid-point was taken as the representative value. Observed IVE at vaccine coverage *x*, was calculated as 1 − *IR*_x_⁄*IR*_0_, where *IR*_x_represents the incidence rate among unvaccinated people in a population with vaccine coverage rate x, and *IR*_0_ denotes the incidence rate in a population with no vaccination. While *IR*_0_ was not provided in the original analysis of the clinical trial, a subsequent dynamic modeling study^3^ provided a plausible value of *IR*_0_, which was used to compute IVE. This approach resulted in five IVE values at 5 vaccine coverage rates.

To model the relationship between IVE and vaccine coverage, we used a Bayesian Beta regression. The expected value of IVE, *Y*, was an expit-transformed function of *η*, which itself was modeled as a linear function of the independent variable, vaccine coverage *X*. The response variable *Y* was assumed to follow a Beta distribution, parameterized by its mean, *μ*, and precision, *ϕ*. Weekly informative priors were assigned to the linear predictor coefficients, *β*, and the precision parameter, *ϕ* > 0, based on the standard recommendations for Bayesian analysis.^4^ The regression model was implemented in the probabilistic programming language Stan,^5^ and the code is available on GitHub.^6^

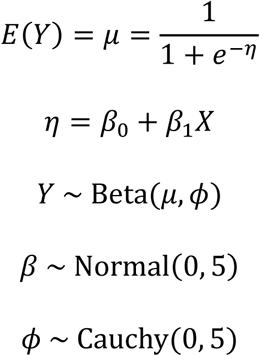

**Figure S1.**
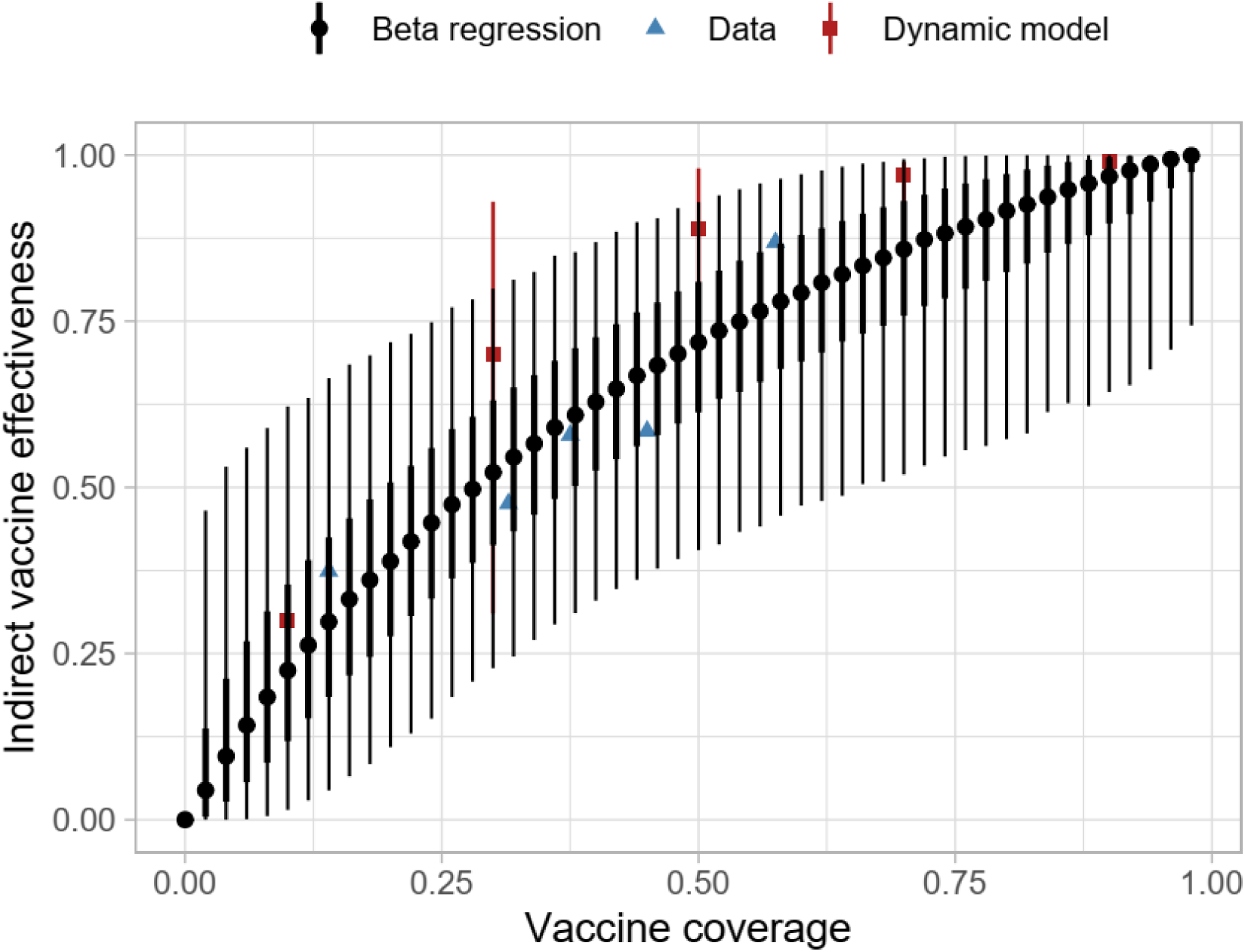
Posterior predicted values for the indirect vaccine effectiveness based on a Bayesian beta regression with cholera incidence data from Matlab, Bangladesh. Dynamic model represents the values from the dynamic model by Longini et al.^3^

## S3. Parameter exploration using Sobol low-discrepancy sampling

To comprehensively explore the effects of key model parameters on cholera outbreak dynamics and the impact of OCV campaigns, we conducted a systematic parameter sampling using the Sobol low-discrepancy sequence.^7,8^ This method provides an efficient way to thoroughly cover complex parameter spaces while minimizing clustering or overrepresentation of specific regions, making it particularly suitable for sensitivity and uncertainty analyses.

We examined a range of plausible values for the following parameters: vaccine efficacy (<5 years and 5+ years), indirect vaccine effectiveness, delay from vaccination and development of partially protective immunity, and campaign duration (days required to complete vaccination in a target population). Different distributions are assumed for each parameter based on the assumption of the estimation and on our best judgement. For the direct vaccine efficacy, we assumed that the log incidence rate ratio (e.g., vaccine efficacy = 1 – incidence rate ratio) as the confidence interval of the original estimates may usually be constructed this way. Indirect vaccine efficacy was drawn from the posterior predictive values of the Bayesian beta regression model we have developed (**S2**). Delay from vaccination and development of partially protective immunity has been assumed to be uniformly distributed due to the paucity of the data. The duration of a vaccination campaign was assumed to follow a truncated normal distribution with the minimum and maximum based on the data and our judgement.

We employed the Sobol low-discrepancy sequence to generate pseudo-random numbers for each parameter, ensuring uniform coverage across their respective distributions. Each Sobol sequence element, ranging from 0 to 1, was transformed into the target parameter space using the inverse cumulative distribution function (CDF) of the specified distribution. The Sobol sequence generation was implemented using the ‘pomp’ R package, with custom routines to map values onto the respective distributions (GitHub^6^). A total of 200 parameter sets were utilized, covering the entire feasible range of values for each parameter.

Validation involved visually inspecting histograms of the generated parameter values to confirm that the transformations were accurate and that the values adhered to the intended distributions.

## S4. Delay in immunogenicity of oral cholera vaccine

We assumed a period of delay before protective immunity develops following vaccination. Studies investigating the immunogenicity of OCV typically measure the rate of seroconversion (considered a proxy for protection), defined as a four-fold or greater increase in antibody/vibriocidal titres compared to baseline titres.^9–17^ However, dosing schedules, follow-up times, and populations varied across studies. For instance, Ng’ombe et al. investigated a two-dose schedule for the Shanchol OCV in Zambia, administering the first and second doses 28 days apart and collecting blood samples at baseline and after day 28 (before second dose), month 6, month 12, month 24, month 30, month 42, and month 48.^9^ They observed a significant rise in vibriocidal antibody titres at day 28, which declined nearly to baseline levels at month 6. Other studies by Mohan et al. (India) and Chowdhury et al. (Bangladesh) investigated a two-dose schedule for Shanchol with doses being administered 14 days apart instead, and blood collections performed at days 0, 14 (before second dose), and 28.^11,12^ Both studies found that vibriocidal antibody titres were significantly increased at day 14 and day 28, compared to baseline, and that this increase was higher after the first dose, compared to the increase following the second dose of vaccine. Charles et al. (Haiti) investigated a two-dose Shanchol schedule administered 28 days apart, with blood collection occurring 7 days post-vaccination for each dose.^13^ They found that each age cohort had a significant vibriocidal antibody response by as early as day 7, compared to baseline, and increased further by day 21. Additionally, O-antigen specific IgA serum responses were also significant at day 7 and 21. We believe that these findings make it reasonable to conclude a time to immunity following immunization of 14 days for modeling purposes. This assumption will be tested via sensitivity analyses using a range of values (from 7 to 28 days) to explore the uncertainty of the parameter.

## S5. Number of workdays lost to patients and caregivers

To estimate the average number of productivity days lost due to cholera illness, we compiled patient-level data from four published studies conducted in sub-Saharan Africa between 2012 and 2017. These studies were drawn from diverse geographical settings including Malawi,^18^ Ghana,^19^ Mozambique,^20^ and Zanzibar.^21^ Two additional studies from Somalia^22^ and Zambia^23^ were reviewed but excluded. The Somalia study reported only the average duration of stay at the cholera treatment center only and did not account for post-discharge recovery while the Zambia study provided only categorical durations without reporting a mean or median, and the upper bound of the categories was undefined. The four included studies used varying definitions of productivity loss, such as self-reported inactivity periods, missed workdays, and illness duration. We treated these as conceptually comparable, interpreting each as reflecting the total number of days of productivity lost per patient, including both hospitalization and recovery periods. Two studies (Malawi,^18^ Ghana^19^) also reported productivity loss from the caregiver’s perspective. Using these data, we calculated a weighted average of days lost per episode, applying applying study-specific sample sizes as weights. This yielded an average of 6.5 days per patient and 3.3 days lost per caregiver per cholera episode.

## S6. Mean age of infection

The mean age of infection was estimated as the weighted mean of infected individuals’ ages, based on outbreak investigations in Sudan,^24^ South Sudan,^25^ Nigeria,^26^ Ethiopia,^27^and Uganda.^28,29^ In the Nigeria study,^26^ the mean age was approximated using the formula, (Q1 + 2*median + Q3) /4, which closely matched the actual mean in a Ugandan outbreak,^30^ where median, Q1, Q3, and mean were all reported. In the Ethiopia study,^27^ age-specific case counts were provided; the midpoint of each age group was used as the representative age. For the oldest age group (>=45 years), we assumed a midpoint of 60 years. Similarly, for the >= 60 age group in the Uganda study,^29^ the midpoint was assumed to be 67.5 years. These choices were both based on an estimated upper age bound of 75 years.

## S7. Vaccine coverage

We explored the vaccine coverage rates ranging from 60 to 95 percent of the population. Vaccine coverage rates were chosen to account for observations from OCV campaigns during 2013-2018^31^ and a recent OCV campaigns in Ethiopia.^32^ In particular, we chose 75% as the most representative value for a single-dose regimen. This value was obtained by first computing the ratio of estimated coverage to the administrative coverage, *r*, and then multiplying *r* with the observed administrative coverage during the first dose, *AC*_1_. *r* was computed the estimated by taking the ratio of estimates to administrative coverage for the at least one dose from the OCV campaigns. (*EC*_≥1_and *AC*_≥1_, respectively). *EC*_≥1_were available in the study and were derived assuming random allocation during vaccination during the first and the second dose. Let *AC*_1_and *AC*_2_be administrative vaccine coverage rates for the first and the second dose, respectively. Then,

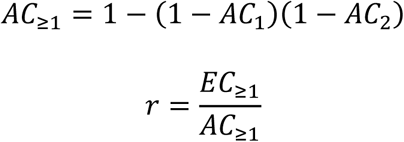

In particular, we used *EC*_≥1_during humanitarian crises or outbreaks (73.2%, and 86.3%, respectively). Finally, we computed the estimated coverage rates for the first dose during humanitarian crises or outbreaks (62.6% and 85.3%) using the following relationship.

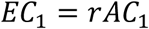

Arithmetic mran of *EC*_1_during humanitarian crises or outbreaks is 74% and used 75% as a convenient proxy value.

## S8. Cost-effective analyses (CEA)

Disability-adjusted life years (DALYs) were computed as:

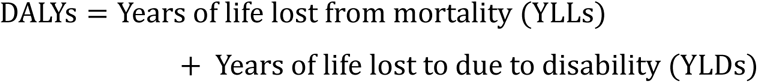

YLLs were computed by multiplying the number of deaths with life-expectancy at the average age of death. Average age of death was derived from the average of infections observed during outbreaks and clinical trials as explained in **S6**. Life expectancy at a given age, mean remaining years of life expected by an individual at that age, was derived from United Nations Life Expectancy at Exact Age for both sexes.^44^ YLDs were computed by multiplying the number of cases with the duration of the disease in years and the disability weight. The duration of disease was assumed to be 2 days and the disability weight was assumed to be 0.188 and 0.247 to account for the moderate and severe cases a based on the disability weights of diarrhea from the 2019 Global Burden of Disease Study,^42^ assuming that reported cases in our data set were moderate or severe cases.

We evaluated cost-effectiveness by calculating the cost per case, death, and DALY averted. Costs related to the cholera vaccine and illness were selected based on recency, generalizability to our modeling data, and quality after a literature review. Prices were adjusted to 2023 USD$ cost-equivalents by converting the costs from the reporting year and inflating them based on the sub-Saharan African average annual consumer price index percent change for each year up until 2023.^45–47^ Consumer prices were obtained from the publicly available International Monetary Fund database.^48^ This was performed using the following equation:

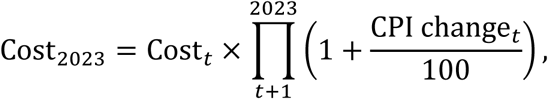

where *t* is the year that the original cost was reported, and CPI change_*t*_ is the consumer price index annual percent change for each year *t*.

The vaccine procurement cost ($1.65) and shipping cost ($0.06) were sourced from UNICEF reports published in 2024, which did not specify the USD year.^49,50^ We assumed these costs were comparable to 2023 and, therefore, left them unadjusted. After adjusting for inflation, cost of vaccine delivery was set to $2.32. Out-of-pocket costs for those hospitalized was set to $109.32 and out-of-pocket costs for outpatient cases was set to $16.06. Costs on the public health system for those hospitalized and outpatient cases was set to $184.97 and $7.99, respectively.

**Figure S2.**
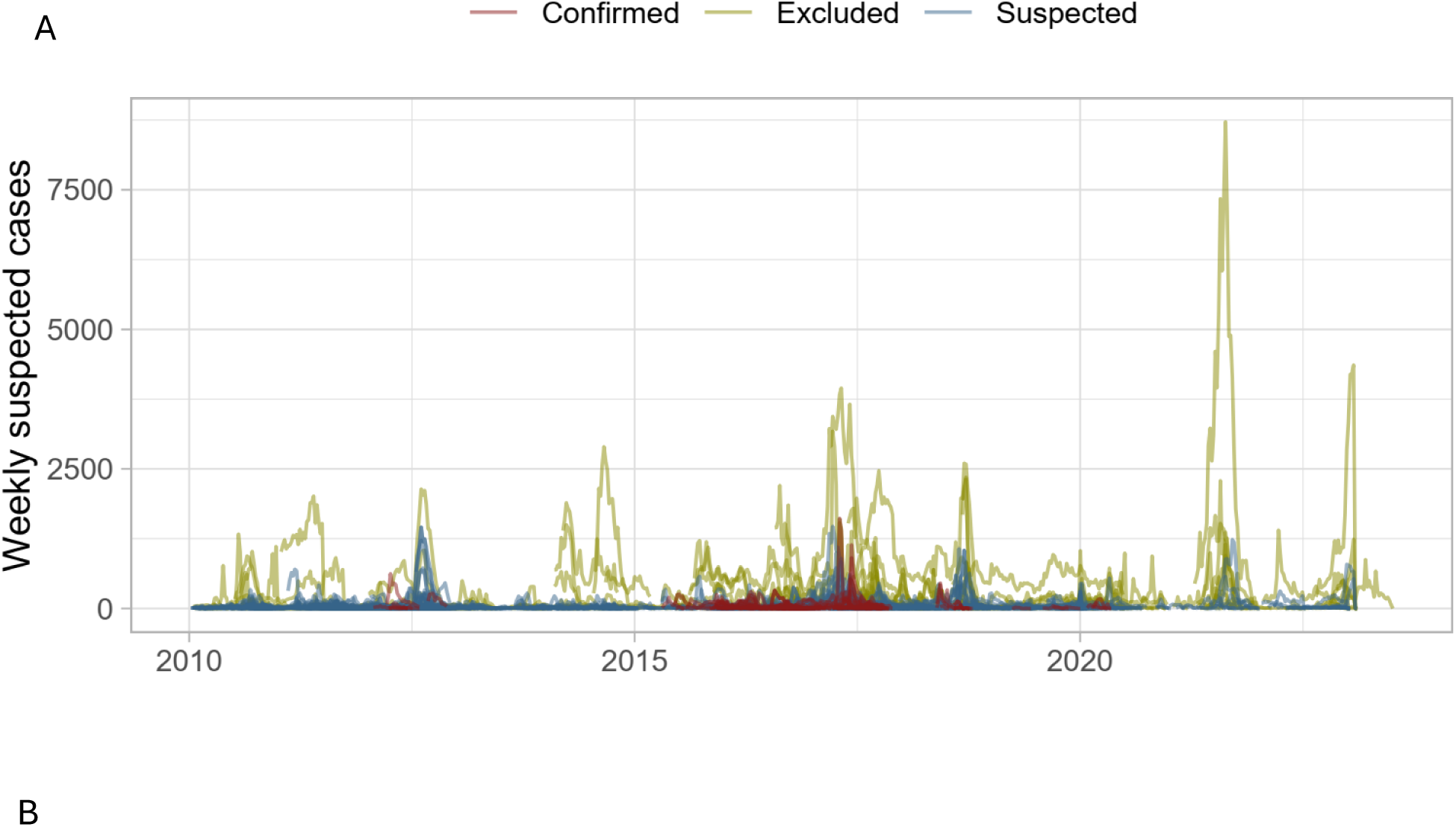
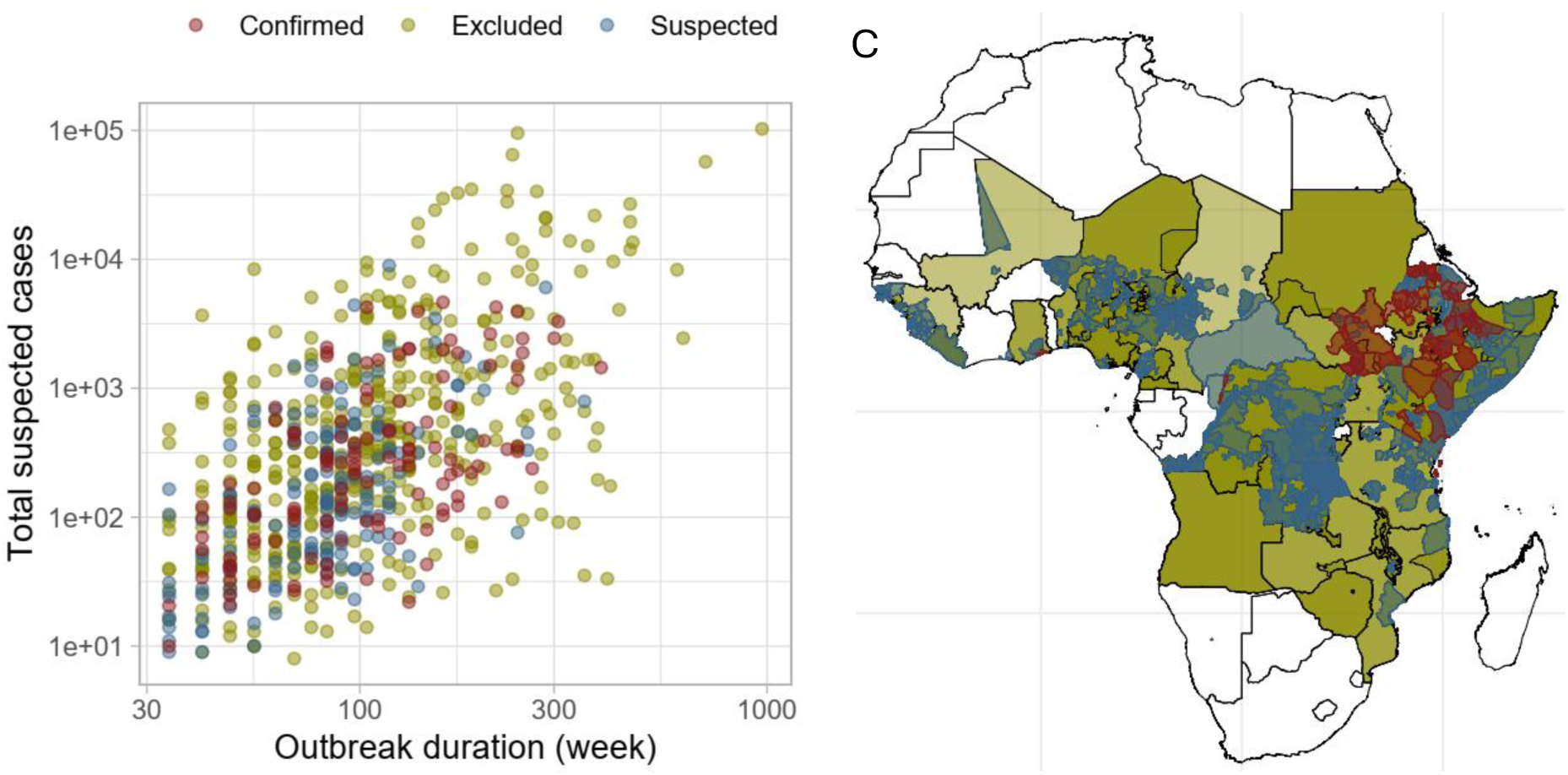
Outbreak data set in the study. Yellow, blue and red colors represent excluded, suspected, and confirmed cholera outbreaks, respectively. (A) weekly suspected cases across time. (B) displays the total number of suspected cases throughout the outbreak against the duration of the outbreak on a log scale for both X and Y axes. (C) shows the geographical location of the outbreak at the administrative unit levels 0,1, and 2.

**Figure S3.**
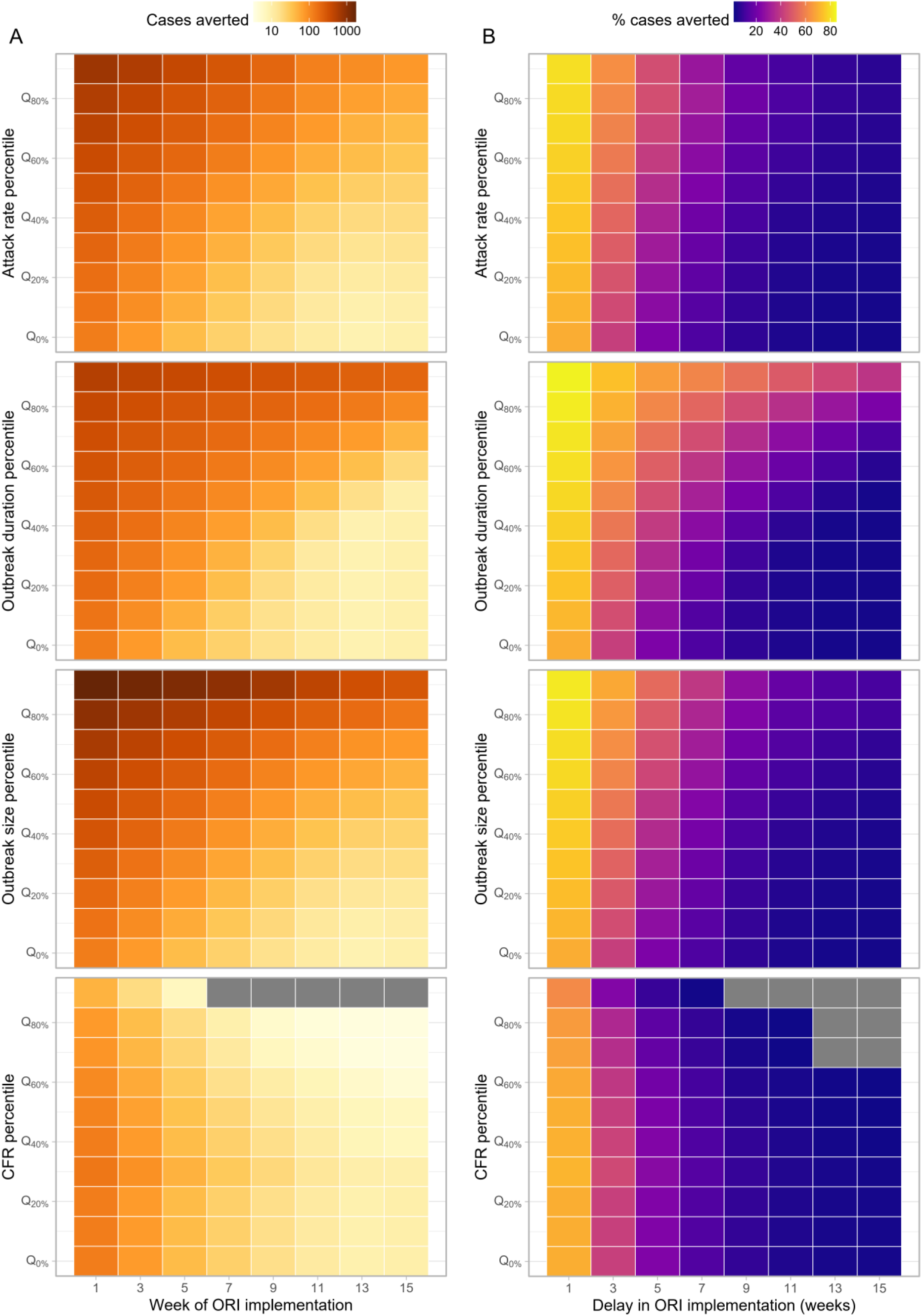
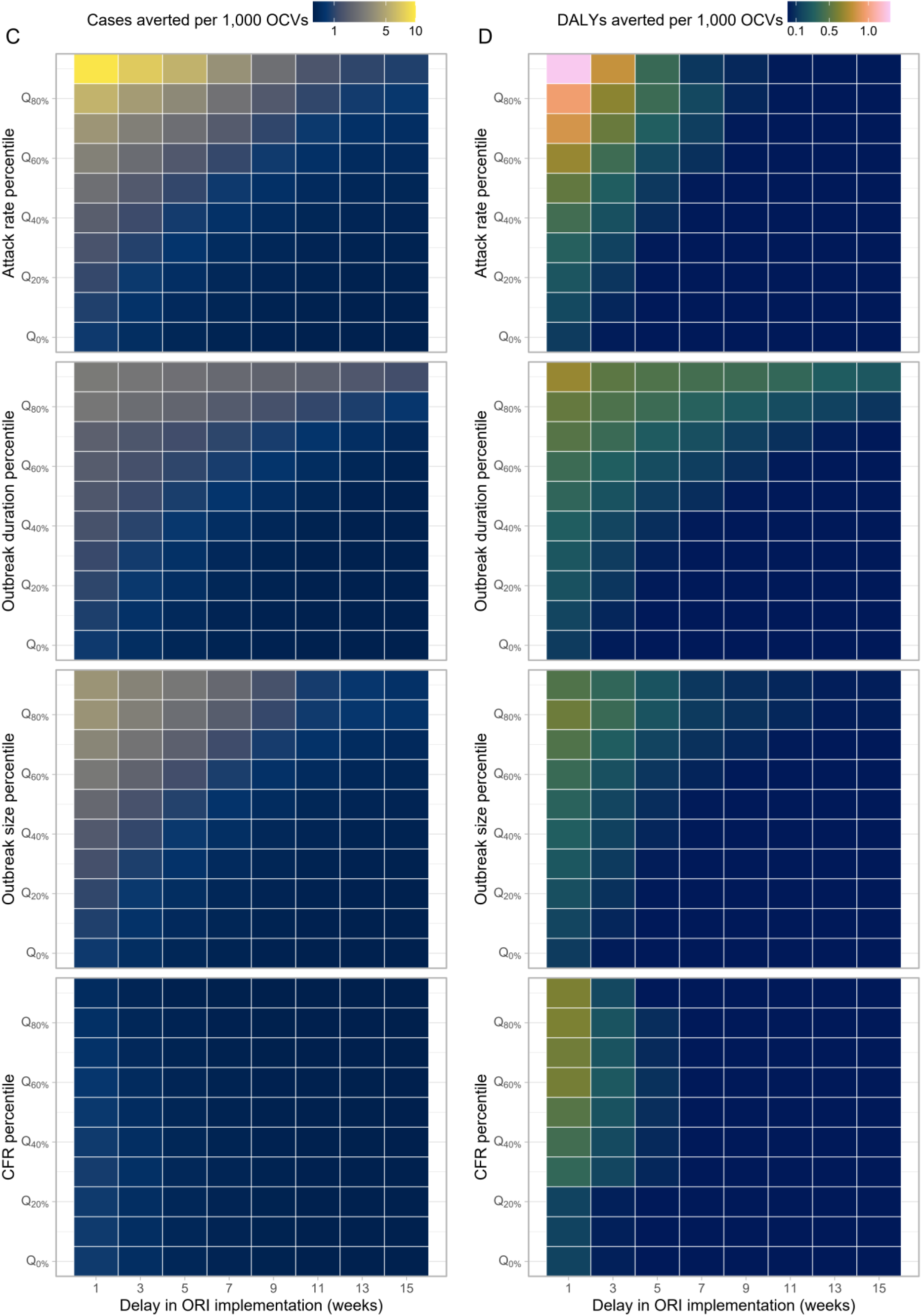
Impact and efficency of outbreak response immunization (ORI) across implementation delays and subsets by attack rate, case fatality ratio, duration, and size. (A) Case averted (B) percentage case averted, (C) case averted per 1,000 OCV and (D) DALY averted per 1,000 OCVs.

**Table S1.** Impact of ORI under the outbreak onset-triggered scenario. Values represent medians with interquartile ranges (IQRs).

| Week of ORI implementation | Vaccine coverage | Outbreaks with ORIs implemented | % case reduction | Case averted | DALY averted | Case averted per 1,000 OCVs | DALY averted per 1,000 OCVs | Cost per case averted | Cost per DALY averted |
| --- | --- | --- | --- | --- | --- | --- | --- | --- | --- |
| 1 | 0.6 | 1,192 (100%) | 64.9 (46.4 - 76.2) | 120 (38 - 347) | 23 (0 - 118) | 0.801 (0.217 - 2.782) | 0.148 (0.001 - 0.787) | 3,914.4 (792.8 - 15,341.3) | 21,079.6 (3,289.9 - 4,025,339) |
| 5 | 0.6 | 1,192 (100%) | 21.1 (2.7 - 44.9) | 36 (3 - 179) | 0 (0 - 43) | 0.229 (0.017 - 1.266) | 0.001 (0 - 0.276) | 8,092.1 (1,515.3 - 34,826.9) | 303,894.4 (6,979.7 - 21,274,967) |
| 9 | 0.6 | 1,084 (90.9%) | 7.2 (0 - 27.4) | 15 (0 - 104) | 0 (0 - 22) | 0.1 (0 - 0.763) | 0 (0 - 0.141) | 11,111.8 (2,145.7 - 51,664.6) | 1,504,166 (9,941.1 - 34,763,429) |
| 13 | 0.6 | 822 (69%) | 3.3 (0 - 19.1) | 8 (0 - 81) | 0 (0 - 20) | 0.054 (0 - 0.571) | 0 (0 - 0.078) | 13,451.2 (2,436.2 - 65,219) | 2,733,219 (11,318.7 - 44,556,414) |
| 1 | 0.75 | 1,192 (100%) | 70 (49.8 - 82.5) | 129 (41 - 375) | 24 (0 - 128) | 0.692 (0.188 - 2.399) | 0.128 (0.001 - 0.679) | 4,661.8 (984.8 - 17,916.7) | 24,770.1 (4,065.8 - 4,679,419) |
| 5 | 0.75 | 1,192 (100%) | 22.8 (3 - 48.4) | 39 (3 - 193) | 0 (0 - 46) | 0.198 (0.015 - 1.093) | 0.001 (0 - 0.238) | 9,486.7 (1,843.7 - 40,532.9) | 354,747.1 (8,338.3 - 24,678,219) |
| 9 | 0.75 | 1,084 (90.9%) | 7.8 (0 - 29.5) | 16 (0 - 112) | 0 (0 - 23) | 0.086 (0 - 0.658) | 0 (0 - 0.122) | 13,042.9 (2,574.7 - 60,025.6) | 1,745,455 (11,816.2 - 40,406,869) |
| 13 | 0.75 | 822 (69%) | 3.6 (0 - 20.7) | 9 (0 - 87) | 0 (0 - 22) | 0.047 (0 - 0.491) | 0 (0 - 0.067) | 15,745.1 (2,912.8 - 75,909.7) | 3,183,230 (13,251.2 - 51,572,816) |
| 1 | 0.9 | 1,192 (100%) | 73.4 (52.1 - 86.3) | 135 (43 - 392) | 25 (0 - 135) | 0.604 (0.164 - 2.088) | 0.111 (0.001 - 0.593) | 5,456.7 (1,195.7 - 20,664.3) | 28,690.9 (4,890.4 - 5,396,109) |
| 5 | 0.9 | 1,192 (100%) | 23.9 (3.1 - 50.6) | 41 (3 - 202) | 0 (0 - 48) | 0.172 (0.013 - 0.953) | 0.001 (0 - 0.207) | 10,975.7 (2,193.8 - 46,544.4) | 410,361.3 (9,840 - 28,345,288) |
| 9 | 0.9 | 1,084<br>(90.9%) | 8.1 (0 -<br>30.9) | 17 (0 - 117) | 0 (0 - 24) | 0.075 (0 -<br>0.573) | 0 (0 - 0.106) | 15,022 (3,036.5 -<br>68,905.7) | 2,004,934<br>(13,899.1 -<br>46,464,569) |
| 13 | 0.9 | 822 (69%) | 3.7 (0 -<br>21.7) | 9 (0 - 91) | 0 (0 - 23) | 0.041 (0 -<br>0.429) | 0 (0 - 0.058) | 18,294.1 (3,427.6 -<br>87,223) | 3,668,705<br>(15,291.5 -<br>59,098,076) |

## S9. Vaccine effectiveness comparison using dynamic models

To assess the robustness of the vaccine impact projections from our static modeling approach, we compared these predictions with those from a dynamic modeling approach. The dynamic model accounts for cholera transmissibility and population immunity, enabling it to capture outbreak-specific variations and feedback loops inherent to epidemic processes—such as the amplification of the force of infection with increasing cases or the mitigating effect of population immunity. While our primary analysis of the static model focused on reactive vaccination scenarios (i.e., outbreak response immunization, ORIs), the comparison between the static and dynamic models was conducted solely for pre-emptive vaccination scenarios. This decision was made after accounting for the limitation in coarse weekly time resolution of the static model and the dynamic model’s limited ability to account for delays in vaccination rollout and subsequent immunological responses, which complicates a direct comparison for ORIs. Pre-emptive vaccination scenarios, on the other hand, provide a more straightforward alignment between the two approaches, allowing us to identify similarities and differences. These differences likely stem from the indirect vaccine effectivness, which is natural in the dynamic model but must be externally specified in the static model.

We employed the standard *Susceptible*-*Exposed*-*Infected*-*Recovered* (*SEIR*) framework in which *E* and *I* states were modeled with two compartments such that the residence time in each compartment follows an Erlang distribution with the shape of 2, rather than the unrealistic exponential distribution (**Figure S3**).

The following is a set of equations to represent the model. The first subscript, *i*, and the second subscripts, *j*, (both 1 or 2) represent the age groups (< 5 yo and 5+ yo) and the *j*th compartments for each state, respectively, except for the *S* state. *S* state and the vaccine efficacy, *χ*, have only one subscript representing the age group.

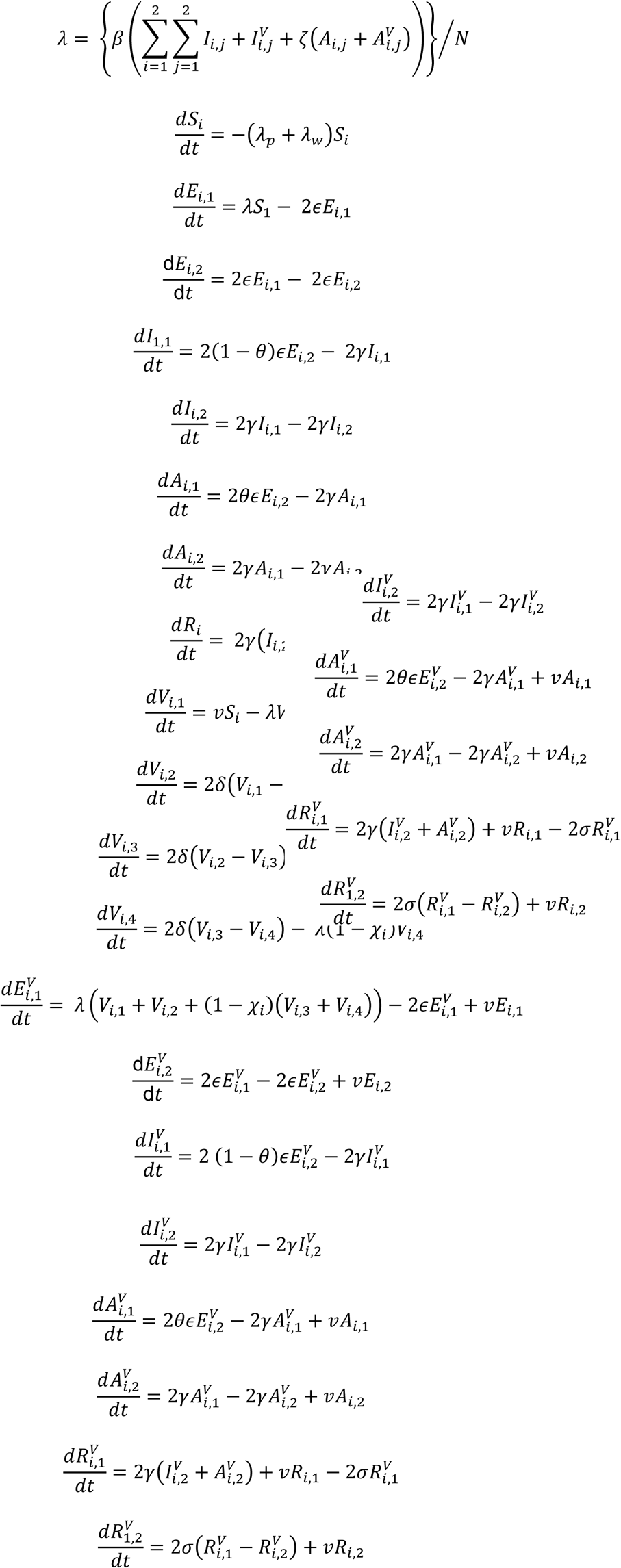

**Figure S5.**
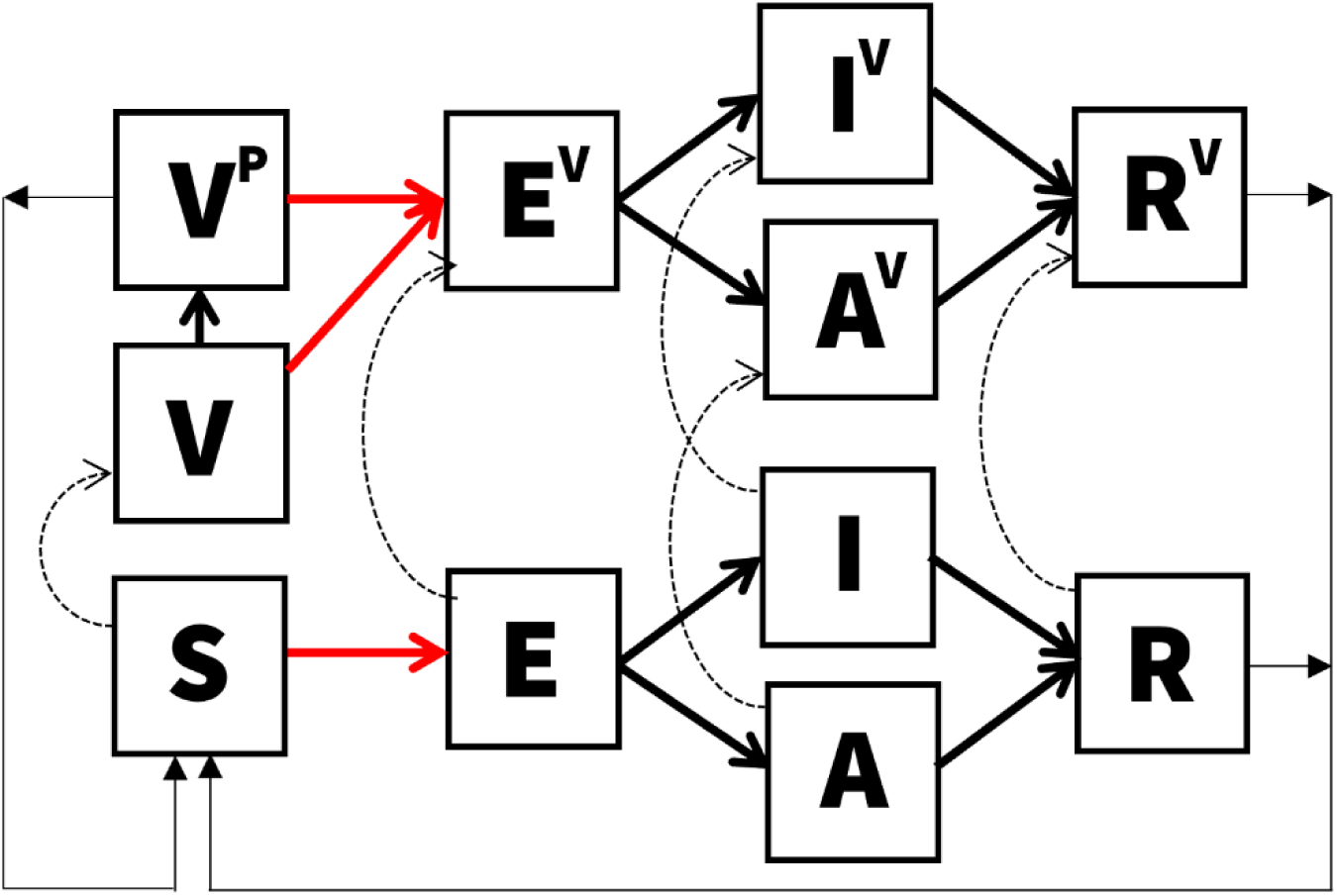
Compartmental flows for the dynamic model. S, E, I, A, R, V represent susceptible, exposed, infectious, asymptomatic, recovered, and vaccinated states. Superscript, P, for V represents those vaccinated and partially protected (rather than vaccinated but fully susceptible V state). Superscript, V, for E, I, A, and R represents vaccinated but infected population. Each compartment is modeled with two sub-compartments representing age groups (<5 yo or 5+ yo) and E, I, A, R, V, and Vp states are additionally modeled with two compartments to model the Erlang-distributed residence times.

The incubation period was modeled using an Erlang distribution. This was achieved by fitting the Erlang distribution to a dataset of 1,000 samples, which were generated through a log-normal distribution with a median of 1.4 days and a dispersion of 1.98, as determined by a systematic literature review.^51^ In our approach, we varied the shape parameter of the Erlang distribution by setting it to 1, 2, 3, and 4. These values represent the potential number of compartments for the incubation state within the ODE model of cholera transmission. The rate parameter was the only variable allowed to be adjusted and was determined through the maximization of the log likelihood. The selection of the optimal shape parameter was based on the comparison of likelihood values, ultimately identifying a shape of 2 and a rate of 1.115044 as the most suitable for our model.

We tried fitting the model to weekly incidence for all 1,406 outbreaks by calibrating four parameters: the proportion of the population that is susceptible in the beginning of the outbreak, *s*_0_, basic reproduction ratio, *R*_0_, initial proportion of people already infected at the start of the outbreak, *i*_0_, and the proportion of effective population size, *n*_0_. The *n*_0_ parameter was introduced to address the possibility that the population experiencing the outbreak is likely to have been a fraction of the population of the administrative unit where the outbreak was reported. Fitting involved assuming that the number of observed cases at a given week, *y*(*t*), follows a Poisson distribution with its mean, *Y*(*t*), and modelled incidence of symptomatic cases. The likelihood function ℒ(θ) of the model, defined by the parameters θ (as shown below), was maximized using the differential evolution method.^52^

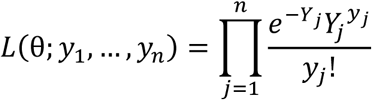

The differential evolution algorithm is a stochastic search algorithm and uses Latin Hypercube sampling as initial parameter values to explore within the bounds and local maxima. We explored a wide range of bounds for parameters to make sure that all reasonable values are examined. Acknowledging that the algorithm may end up in a local minimum by chance, we ran the algorithm with five different random seeds and chose the values that maximized the log likelihood that the algorithm reached at least twice.

After reviewing the fitting results, we selected 341 outbreaks where the model captured the dynamics of the outbreak sufficiently well by excluding fits where a) modeled and observed weekly incidences diverged upon visual inspection, b) parameter estimates were too close (0.1%) to the bounds of parameter inputs, raising concerns that the estimated parameter values were not robust, and c) the number of data points was smaller than 8, corresponding to twice the number of data points. While the number of parameters to be estimated is four, we consider the possibility that the number of data points may need to be much larger to inform the parameter values in practice. The dynamic model was implemented in Julia language^53^ and codes used are available on GitHub.^6^

**Figure S6.**
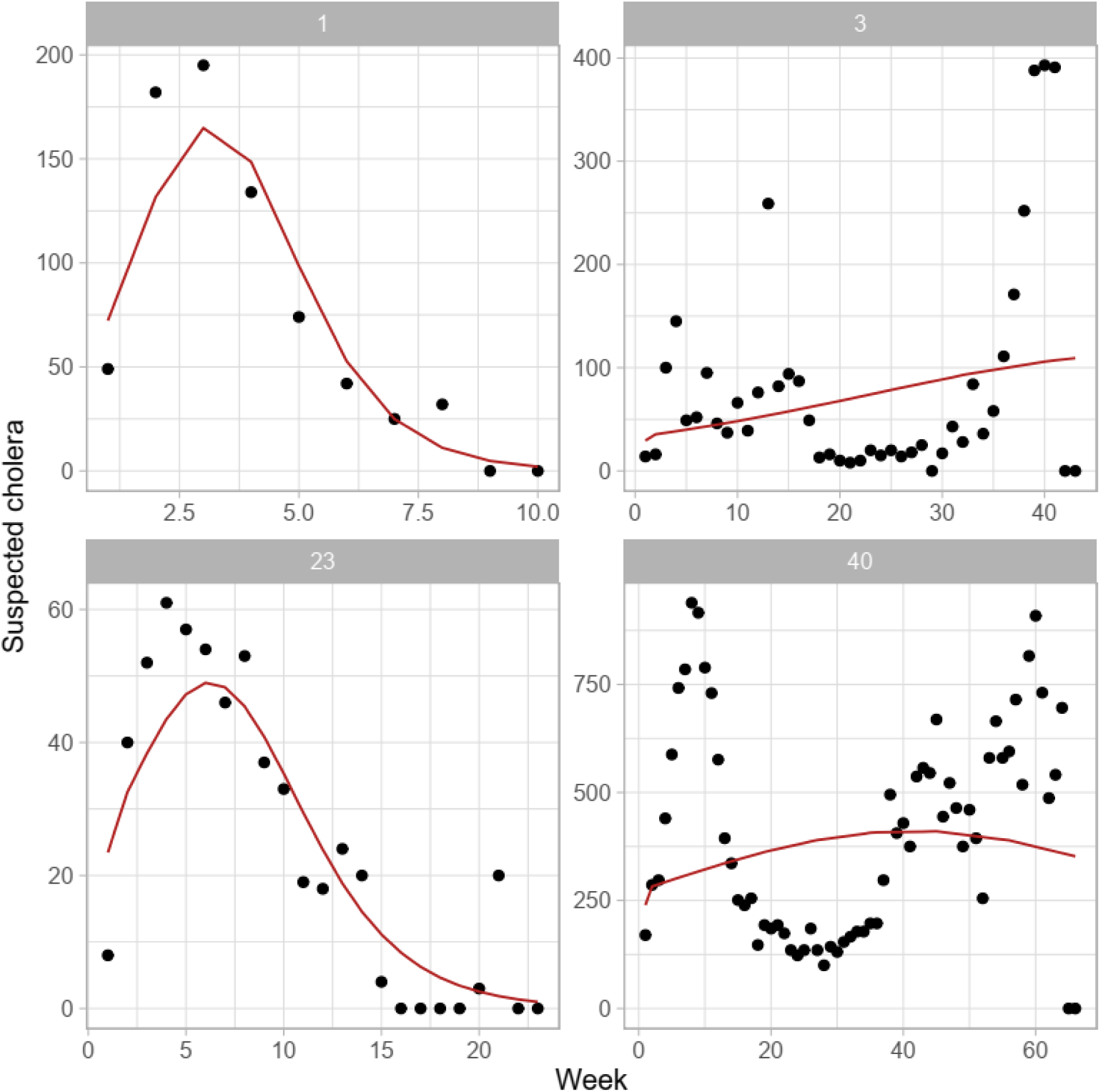
Comparison of weekly incidence between the dynamic model and the observed data. Numbers at the top indicate IDs assigned to each outbreak. For outbreaks with IDs 1 and 23, the dynamic model (red lines) successfully captured the observed data (black dots). Conversely, outbreaks with IDs 3 and 40 exhibited complex patterns that the dynamic model failed to accurately represent.

**Figure S7.**
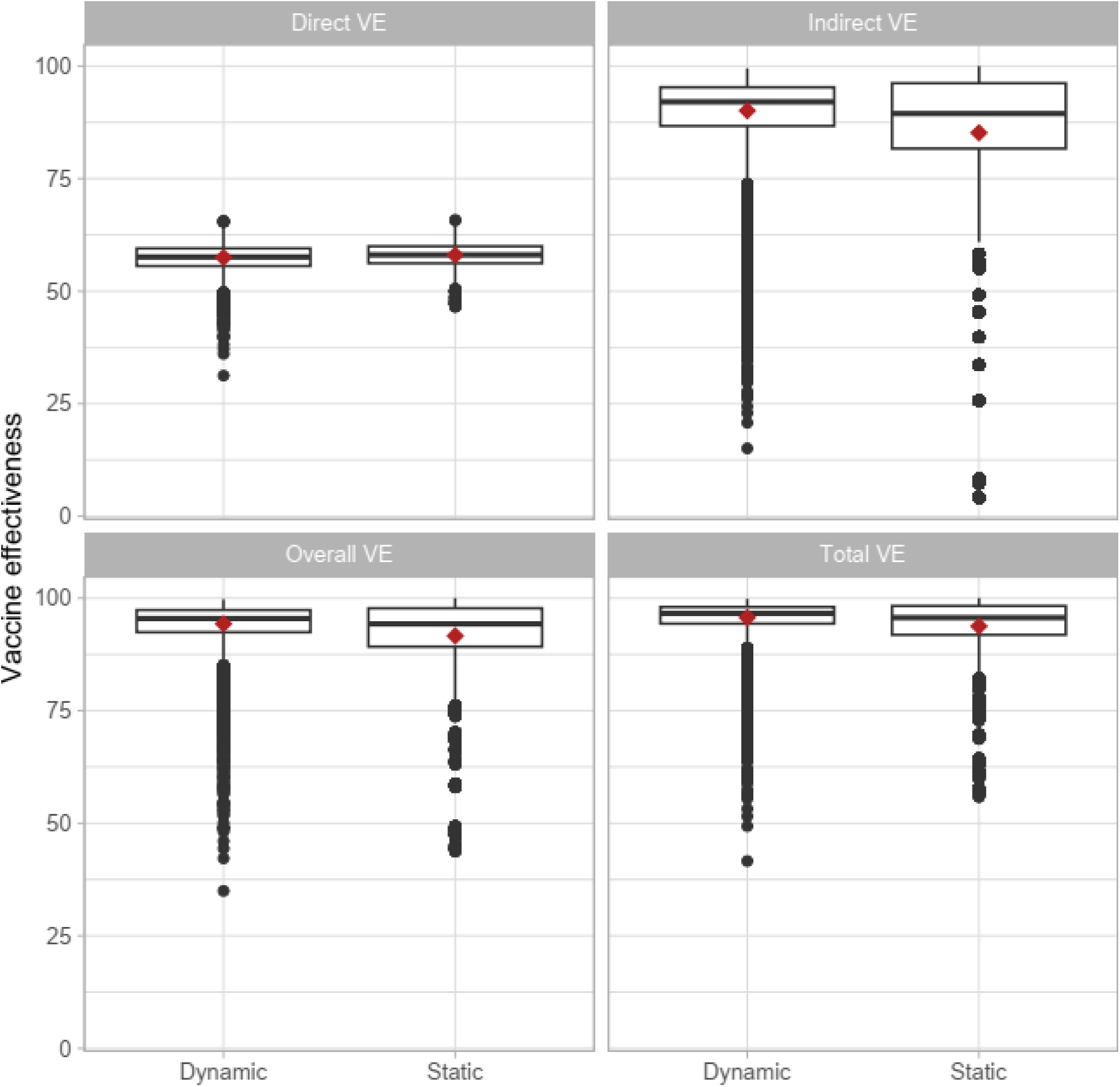
Direct, indirect, total, and overall vaccine effectiveness in the dynamic and static models. Vaccine effectiveness was measured under the pre-emptive vaccination scenario where a proportion of the individuals, determined by the vaccine coverage rate, were in the compartments of individuals who acquired vaccine-derived protection, *V*^*P*^(**Figure S3**), in the beginning of the simulation.

**Table S3.** Impact, efficiency, and cost effectiveness of ORI under the outbreak onset-triggered scenario for varying delays to stop ORI after outbreak end and varying delays in ORI implementation after outbreak onset. Values represent medians with interquartile ranges (IQRs).

| Week of ORI implementation | Delay to stop ORI after outbreak end (weeks) | Outbreaks with ORIs implemented | % case reduction | Case averted | DALY averted | Case averted per 1,000 OCVs | DALY averted per 1,000 OCVs | Cost per case averted | Cost per DALY averted |
| --- | --- | --- | --- | --- | --- | --- | --- | --- | --- |
| 1 | 1 | 1,192 (100%) | 70 (49.8 - 82.5) | 129 (41 - 375) | 24 (0 - 128) | 0.692 (0.188 - 2.399) | 0.128 (0.001 - 0.679) | 4,661.8 (984.8 - 17,916.7) | 24,770.1 (4,065.8 - 4,679,419) |
| 1 | 3 | 1,192 (100%) | 70 (49.8 - 82.5) | 129 (41 - 375) | 24 (0 - 128) | 0.692 (0.188 - 2.399) | 0.128 (0.001 - 0.679) | 4,661.8 (984.8 - 17,916.7) | 24,770.1 (4,065.8 - 4,679,419) |
| 1 | 5 | 1,192 (100%) | 70 (49.8 - 82.5) | 129 (41 - 375) | 24 (0 - 128) | 0.692 (0.188 - 2.399) | 0.128 (0.001 - 0.679) | 4,661.8 (984.8 - 17,916.7) | 24,770.1 (4,065.8 - 4,679,419) |
| 1 | 7 | 1,192 (100%) | 70 (49.8 - 82.5) | 129 (41 - 375) | 24 (0 - 128) | 0.692 (0.188 - 2.399) | 0.128 (0.001 - 0.679) | 4,661.8 (984.8 - 17,916.7) | 24,770.1 (4,065.8 - 4,679,419) |
| 5 | 1 | 1,192 (100%) | 24 (4.2 - 49.1) | 42 (4 - 202) | 0 (0 - 47) | 0.215 (0.022 - 1.143) | 0.001 (0 - 0.25) | 9,486.7 (1,843.7 - 40,532.9) | 354,747.1 (8,338.3 - 24,678,219) |
| 5 | 3 | 1,192 (100%) | 22.4 (2.7 - 48.2) | 38 (3 - 190) | 0 (0 - 46) | 0.192 (0.012 - 1.077) | 0.001 (0 - 0.234) | 9,486.7 (1,843.7 - 40,532.9) | 354,747.1 (8,338.3 - 24,678,219) |
| 5 | 5 | 1,192 (100%) | 22.4 (2.7 - 48.2) | 38 (3 - 190) | 0 (0 - 46) | 0.192 (0.012 - 1.077) | 0.001 (0 - 0.234) | 9,486.7 (1,843.7 - 40,532.9) | 354,747.1 (8,338.3 - 24,678,219) |
| 5 | 7 | 1,192 (100%) | 22.4 (2.7 - 48.2) | 38 (3 - 190) | 0 (0 - 46) | 0.192 (0.012 - 1.077) | 0.001 (0 - 0.234) | 9,486.7 (1,843.7 - 40,532.9) | 354,747.1 (8,338.3 - 24,678,219) |
| 9 | 1 | 1,017<br>(85.3%) | 9.8 (0.2 -<br>31.7) | 22 (1 - 128) | 0 (0 - 24) | 0.112 (0.001<br>- 0.764) | 0 (0 - 0.152) | 13,042.9 (2,574.7 -<br>60,025.6) | 1,745,455<br>(11,816.2 -<br>40,406,869) |
| 9 | 3 | 1,136<br>(95.3%) | 5.8 (0 -<br>27.4) | 12 (0 - 101) | 0 (0 - 23) | 0.063 (0 -<br>0.57) | 0 (0 - 0.094) | 13,042.9 (2,574.7 -<br>60,025.6) | 1,745,455<br>(11,816.2 -<br>40,406,869) |
| 9 | 5 | 1,192<br>(100%) | 3.4 (0 -<br>24) | 6 (0 - 80) | 0 (0 - 21) | 0.03 (0 -<br>0.456) | 0 (0 - 0.053) | 13,042.9 (2,574.7 -<br>60,025.6) | 1,745,455<br>(11,816.2 -<br>40,406,869) |
| 9 | 7 | 1,192<br>(100%) | 2.7 (0 -<br>22.9) | 5 (0 - 74) | 0 (0 - 20) | 0.021 (0 -<br>0.422) | 0 (0 - 0.036) | 13,042.9 (2,574.7 -<br>60,025.6) | 1,745,455<br>(11,816.2 -<br>40,406,869) |
| 13 | 1 | 765<br>(64.2%) | 5.7 (0 -<br>23.4) | 15 (0 - 109) | 0 (0 - 23) | 0.078 (0 -<br>0.605) | 0 (0 - 0.112) | 15,745.1 (2,912.8 -<br>75,909.7) | 3,183,230<br>(13,251.2 -<br>51,572,816) |
| 13 | 3 | 890<br>(74.7%) | 2.1 (0 -<br>18) | 5 (0 - 72) | 0 (0 - 16) | 0.026 (0 -<br>0.387) | 0 (0 - 0.021) | 15,745.1 (2,912.8 -<br>75,909.7) | 3,183,230<br>(13,251.2 -<br>51,572,816) |
| 13 | 5 | 1,017<br>(85.3%) | 0.3 (0 -<br>13.9) | 1 (0 - 49) | 0 (0 - 0) | 0.004 (0 -<br>0.255) | 0 (0 - 0.001) | 15,745.1 (2,912.8 -<br>75,909.7) | 3,183,230<br>(13,251.2 -<br>51,572,816) |
| 13 | 7 | 1,136<br>(95.3%) | 0 (0 -<br>10.8) | 0 (0 - 34) | 0 (0 - 0) | 0 (0 - 0.179) | 0 (0 - 0.001) | 15,745.1 (2,912.8 -<br>75,909.7) | 3,183,230<br>(13,251.2 -<br>51,572,816) |

**Table S3.** Impact of ORI under the outbreak onset-triggered scenario based on the alternative outbreak dataset. Values represent medians with interquartile ranges (IQRs)..

| Week of ORI implementation | Outbreaks with ORIs implemented | % case reduction | Case averted | DALY averted | Case averted per 1,000 OCVs | DALY averted per 1,000 OCVs | Cost per case averted | Cost per DALY averted |
| --- | --- | --- | --- | --- | --- | --- | --- | --- |
| 1 | 1,124 (100%) | 69.9 (49.3 - 82.6) | 120 (38 - 347) | 31 (0 - 135) | 0.623 (0.161 - 2.102) | 0.138 (0.001 - 0.731) | 4,973.2 (1,070.9 - 19,817) | 23,023.9 (3,553.4 - 5,285,194) |
| 5 | 1,124 (100%) | 23.7 (2.8 - 49.7) | 38 (3 - 184) | 0 (0 - 47) | 0.183 (0.011 - 1.033) | 0.001 (0 - 0.287) | 9,525.9 (1,876.6 - 41,559.9) | 150,094.9 (6,744 - 25,584,304) |
| 9 | 1,019 (90.7%) | 9.5 (0 - 31.8) | 18 (0 - 117) | 0 (0 - 25) | 0.092 (0 - 0.667) | 0 (0 - 0.167) | 12,314.7 (2,520.3 - 56,835.9) | 701,831.4 (8,897.5 - 38,101,950) |
| 13 | 776 (69%) | 5.1 (0 - 23) | 12 (0 - 96) | 0 (0 - 23) | 0.06 (0 - 0.536) | 0 (0 - 0.121) | 15,779.6 (2,897.6 - 75,403.2) | 2,578,844 (11,763.9 - 50,219,682) |

